# Feature overlap in models of sustained attention and autism symptoms

**DOI:** 10.64898/2026.04.01.26349372

**Authors:** Corey Horien, Francesca Mandino, Anna Corriveau, Abigail S. Greene, David O’Connor, Xilin Shen, Arielle S. Keller, Erica B. Baller, Marvin M. Chun, Emily S. Finn, Katarzyna Chawarska, Evelyn M.R. Lake, Dustin Scheinost, Theodore D. Satterthwaite, Monica D. Rosenberg, R. Todd Constable

## Abstract

Sustained attention is an important neurobiological process impaired in multiple neurodevelopmental disorders, including attention-deficit/hyperactivity disorder (ADHD) and autism. Connectome-based predictive models (CPMs) of sustained attention function and autism symptoms generalize across samples, but whether their features are shared or phenotype-specific is unknown. We tested within-phenotype consistency and across-phenotype specificity in 5 published CPMs predicting autism symptoms or sustained attention performance in youth and adults, equating model size (top 1,000 positively and negatively predictive edges per model). No edge or network pair was shared by all 5 models. Same-phenotype models were nonetheless consistent: the whole-brain pattern of edge-behavior associations correlated in all 4 same-phenotype dataset pairs (r > 0.13, P < 0.005). Features were also largely phenotype-specific: only 1 of 6 cross-phenotype pairs correlated significantly (negatively), and 2 models built to predict autism symptoms vs. attention in the same dataset shared no more edges than chance. Finally, representational similarity analysis revealed that scan type (rest vs. task), participant age, and autism diagnosis did not explain similarity in edge-behavior associations across datasets. Together, these results suggest that feature sets are reproducible within, but distinct across, phenotypes, consistent with autism symptoms and sustained attention being related but separable.

**Highlights:**

- We interrogated five previously published, transdiagnostic models of sustained attention and autistic phenotypes
- Functional connectome-based models shared edges and networks when phenotypes were similar
- Models predicting attention phenotypes shared more similar features with each other than models predicting autism symptoms and vice-versa
- Age, functional run type, and diagnosis did not affect model similarity

## Introduction

The ability to pay attention is central to human experience. Whether focusing during conversations, monitoring other drivers on the highway, or searching for groceries in a crowded supermarket aisle, the capacity to sustain attention influences nearly every waking moment. Perhaps in part because of its ubiquity, ‘sustained attention’ is a broad construct with poorly understood brain correlates (Rosenberg et al. 2016). There has been significant interest in the field of cognitive and clinical neuroscience in identifying fMRI-based patterns associated with the ability to sustain attention (e.g. Bonnelle et al. 2011; Thompson et al. 2013; Kessler et al. 2016; Rosenberg et al. 2016; Rohr et al. 2017; O’Halloran et al. 2018; Cai et al. 2021; Kardan et al. 2022; Seeburger et al. 2024; Corriveau et al. 2025).

Beyond its role in daily life, sustained attention is important in neurology and psychiatry. Sustained attention is associated with numerous clinical conditions, including dyslexia in youth (Facoetti et al. 2000; Shaywitz and Shaywitz 2008) and delirium in medically ill patients (Ramírez and Schoo, 2022). Difficulties with sustained attention also play an important role in neurodevelopmental disorders, such as attention-deficit/hyperactivity disorder (ADHD) and autism spectrum disorder (Garretson et al. 1990) (hereafter, ‘autism’). It is estimated that approximately 8% of youth worldwide (Ayano et al. 2023) have been diagnosed with ADHD, with that number reaching 10.5% of youth in the United States (Danielson et al. 2024). Approximately 3.2% of eight-year-olds in the US carry an autism diagnosis (Shaw et al. 2025); worldwide, approximately 1 in 127 individuals have been diagnosed (Santomauro et al. 2025).

Although they are distinct diagnoses, there is evidence that ADHD and autism have a number of similarities. For instance, patients in both conditions share a number of behavioral features, including sensory processing issues (Lau-Zhu et al. 2019), executive dysfunction (Corbett et al. 2009), irritability (Brereton et al. 2006), and metacognitive difficulties, such as performance monitoring (Lau-Zhu et al. 2019). Furthermore, difficulties sustaining attention have been reported in both conditions (Garretson et al. 1990; Corbett and Constantine 2006), but the exact behavioral and brain findings are heterogeneous (Johnson et al. 2007; Huang-Pollock et al. 2012; Christakou et al. 2013; Lukito et al. 2020; Cortese et al. 2021; Tamon et al. 2024; Zhou et al. 2026). As an example, when ADHD and autistic participants performed an fMRI vigilance task with increasing sustained attention demands, similar brain areas were recruited including bilateral striato–thalamic regions, left dorsolateral prefrontal cortex (DLPFC), and superior parietal cortex (Christakou et al. 2013). Differences were appreciated as well, however, with autistic participants showing more cerebellar activation. ADHD participants showed less activation of DLPFC, which was associated with worse performance on the task. The differences in brain areas involved in the task led to differential performance, with the ADHD participants exhibiting more sustained attention difficulties (longer response times) (Christakou et al. 2013). Differences in subcortical arousal systems (e.g., Zhou et al. 2026) have been suggested to account for the differences in sustained attention performance in ADHD individuals compared to autistic individuals (Johnson *et al*. 2007), specifically the increased response times typically observed in sustained attention tasks. This is in line with a recent report (Kay et al. 2025), using precision functional mapping methods (Gordon et al. 2017), suggesting that stimulants often prescribed for ADHD seem to affect arousal-related brain networks rather than attention networks (Rubia et al. 2009).

Similar to Christakou et al. (2013), other studies using resting-state fMRI to compare individuals with ADHD to individuals with autism have reported similar patterns of shared brain areas, along with condition-specific differences. In one study (Di Martino et al. 2013), the precuneus exhibited increased degree centrality in both conditions. Nevertheless, differences were found across diagnoses, with increased degree centrality in right striatum and pallidum in ADHD, whereas autism participants exhibited increases in bilateral temporal lobes. More recent work using Bayesian modelling methods (Kernbach et al. 2018) has identified shared resting-state patterns among ADHD and autistic participants involving the default mode, dorsal attention, and the salience networks, with three distinct connectivity factors observed. Factor 1 showed greater expression in those with ADHD, while factor 2 showed greater expression in those with autism, and factor 3 showed no clear relationship with either diagnosis.

Given associations between sustained attention and autism, previous work has focused on the generalizability of predictive models related to attention and autism. For example, a functional connectome-based, cross-validated model defined to predict autistic phenotypes (social responsiveness scale (SRS) scores; Constantino et al. 2003) can successfully generalize in external samples to predict ADHD traits (Lake et al. 2019). Similarly, a model predictive of Autism Diagnostic Observation Schedule-2 (ADOS-2) scores (Lord et al. 2012) generalized to predict sustained attention performance in an independent dataset (Horien et al. 2026), suggesting that there appears to be common functional features associated with autism and attention. In other work, sustained attention functional connectome-based models generated in adults generalized to predict ADHD symptoms in children (Rosenberg et al. 2016). Furthermore, models built to predict sustained attention in youth with autism can generalize to predict sustained attention in neurotypical adults (Horien et al. 2023), and models built to predict visual attention can generalize to auditory attention (Corriveau et al. 2025), indicating there might be a common neurobiological architecture of sustained attention in predictive models.

However, the degree to which these same models are neurobiologically specific has not been formally studied, and it is unclear if models predictive of autistic and attention phenotypes rely on the same underlying brain networks. In particular, despite being used to predict similar phenotypes, the degree to which the models rely on the same functional features has not been investigated. To address the issue of specificity, we interrogated data from five previously published models of sustained attention and autism (Figure 1; Table 1) (Rosenberg et al. 2016; Lake et al. 2019; Horien et al. 2023; Corriveau et al. 2025; Horien et al. 2026). We have three main aims in this work: One, to determine if there is neurobiological overlap in the functional features (edges and networks) in all of the models. Two, to evaluate feature overlap in models predicting the same phenotypes. Three, to conduct exploratory analyses to assess factors influencing similarity in the brain-behavior models, such as age, diagnosis, and other dataset variables.

**Figure 1.**
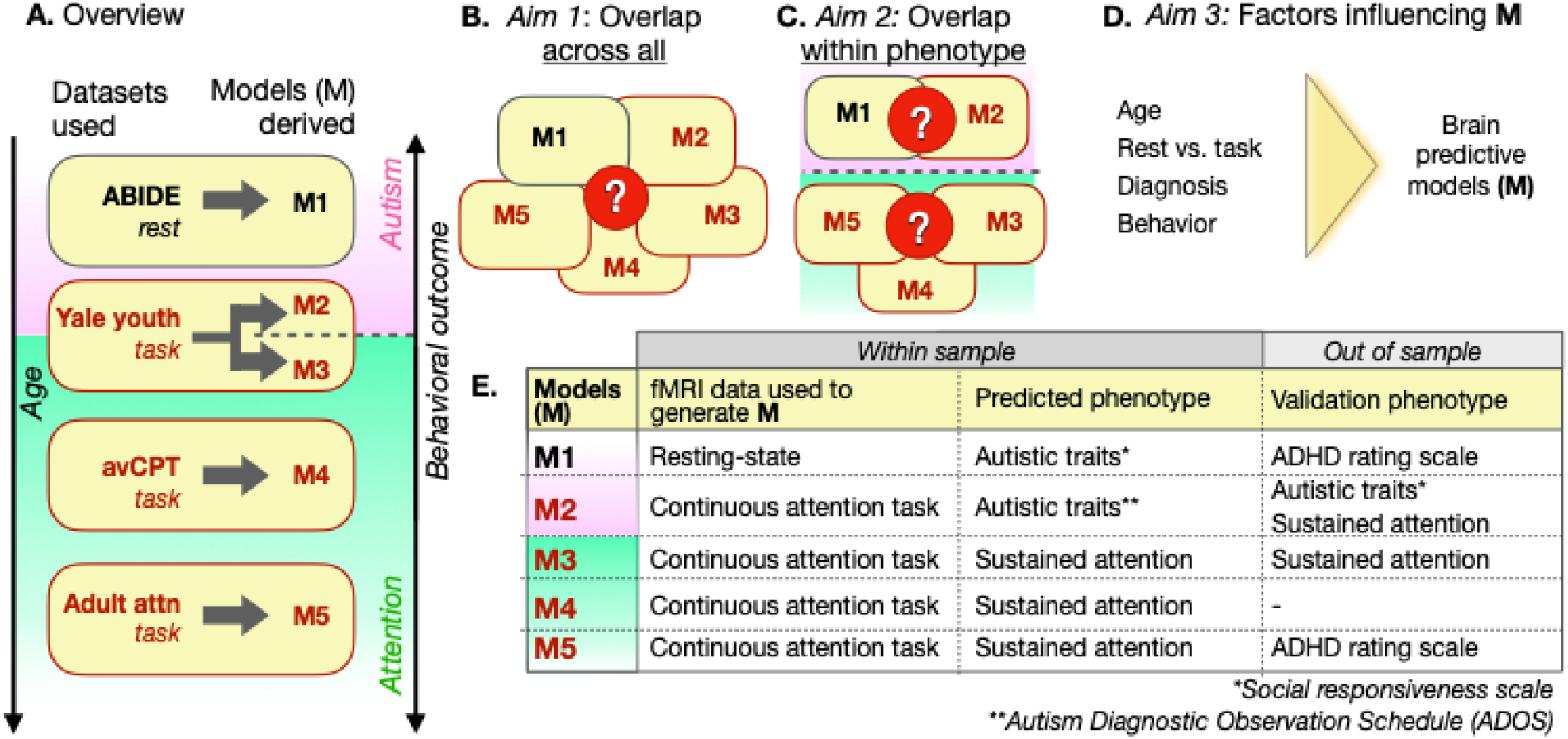
Overview of the current paper. A) Study design. The original sample from which each model was derived is shown in the oval, with the ovals arranged by increasing age as indicated by the arrow to the left. The fMRI data type (resting-state or task) used to generate the models are shown directly under the dataset name. The ABIDE and Yale youth datasets comprised participants with and without autism (indicated by the pink gradient shown around the ovals, with darker pink indicating a higher proportion of participants with autism). The adult attention sample and avCPT sample comprised neurotypical participants. Autism phenotypes were used to generate models M1 and M2 (SRS total scores and ADOS total calibrated severity scores, respectively). Attention phenotypes were used to generate models M3, M4, and M5 (gradCPT task performance). Note that the same parent sample (the Yale youth sample) was used to generate the M2 and M3 models. B) The first aim of the paper was to determine if there was neurobiological overlap across all models. We hypothesized that we could detect overlap, because models trained to predict autism symptoms contain information about sustained attention (and vice-versa). That is, variance in autistic traits correlates with sustained attention (Corbett and Constantine 2006; Lin et al. 2023; Lake et al. 2019; Horien et al. 2023; Horien et al. 2026). C) The second aim was to determine overlap when models predicted congruent phenotypes (autism or sustained attention, top and bottom Venn diagrams, respectively). D) The third aim of the paper is to investigate the impact of different dataset features on the brain-behavior relationships, including age, functional run type (rest vs. task data), diagnosis, and behavioral measure predicted. E) Specifying how each model relates to attention. The leftmost column specifies the model (M1-M5). Each column specifies a model parameter of interest and how it relates to attention, either within-sample to generate the original model or out of sample to externally test the original model. ABIDE, autism brain imaging data exchange; Attn, attention; avCPT, audio-visual continuous performance task. M, model. Alt text: Schematic showing an overview of the study, with a brief summary of the models involved. Also included are the aims of the study, with Venn Diagrams and a semi-flow chart, as well as a table schematic showing brief details of each predictive model.

**Table 1.**
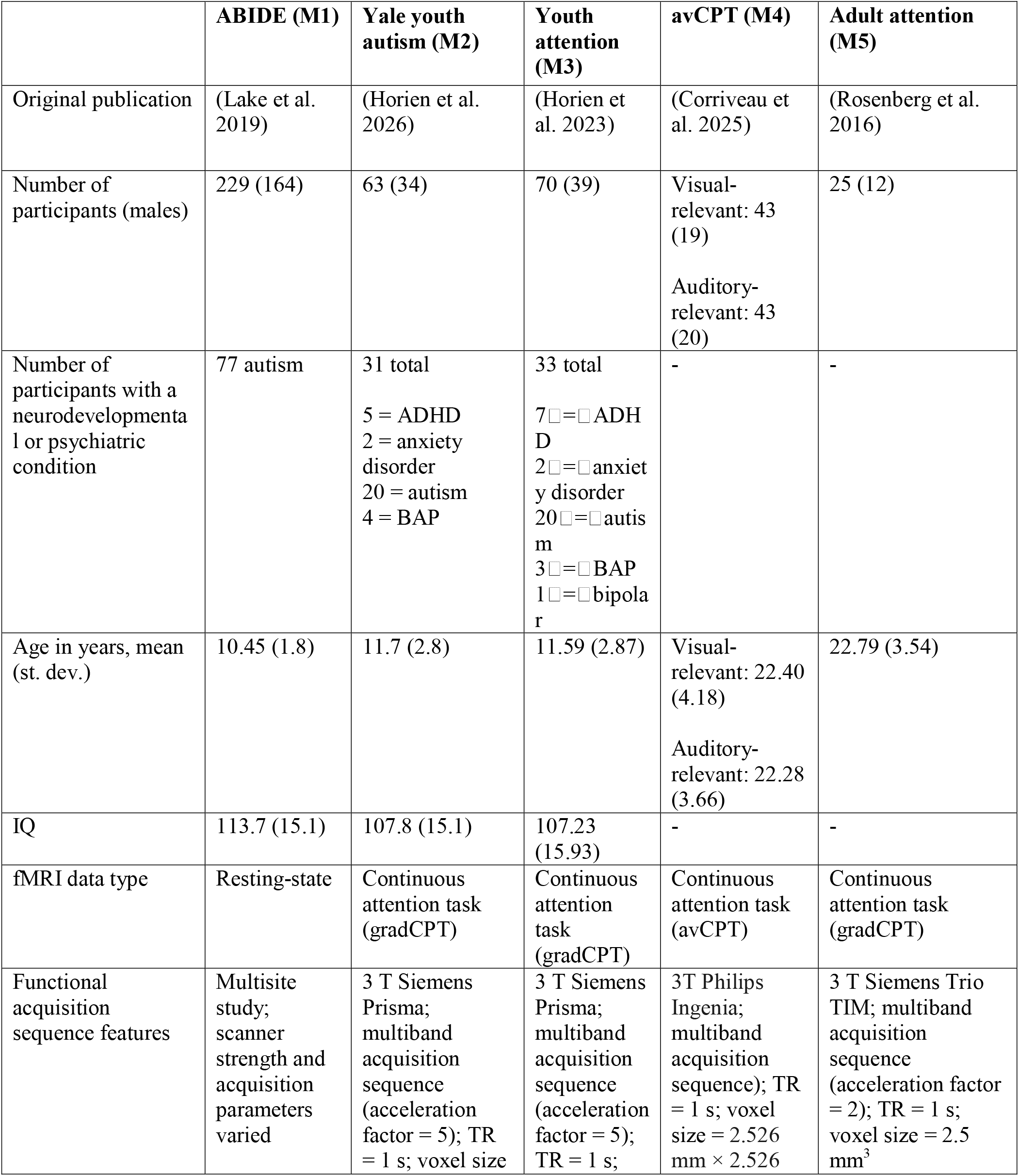
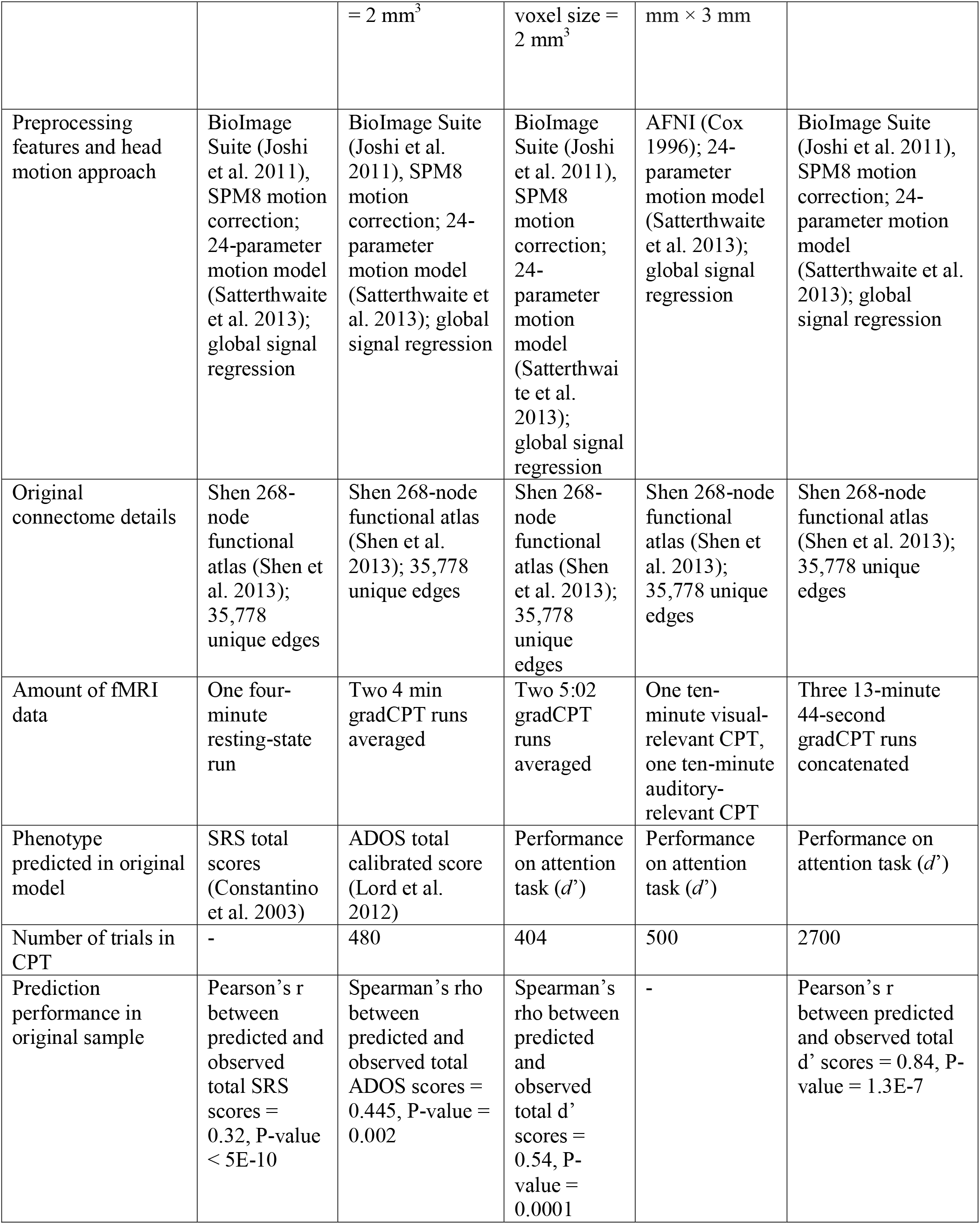
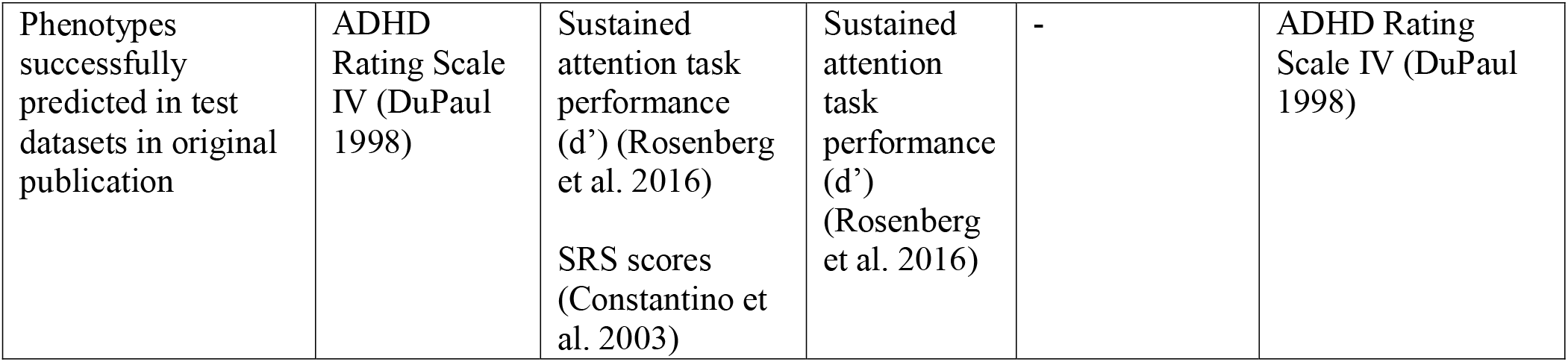
Demographics and modelling parameters of original datasets used to generate predictive models. ABIDE, autism brain imaging data exchange; ADHD, attention-deficit/hyperactivity disorder; AFNI, Analysis of Functional NeuroImages; ADOS, Autism Diagnostic Observation Schedule; avCPT, audio-visual continuous performance task; BAP, broader autism phenotype; CPM, connectome-based predictive modelling; CPT, continuous performance task; gradCPT, gradual-onset continuous performance task; IQ, intelligence quotient; SPM, Statistical Parametric Mapping; s, second; SRS, social responsiveness scale; TR, repetition time.

|  | **ABIDE (M1)** | **Yale youth autism (M2)** | **Youth attention (M3)** | **avCPT (M4)** | **Adult attention (M5)** |
|---|---|---|---|---|---|
| Original publication | (Lake et al. 2019) | (Horien et al. 2026) | (Horien et al. 2023) | (Corriveau et al. 2025) | (Rosenberg et al. 2016) |
| Number of participants (males) | 229 (164) | 63 (34) | 70 (39) | Visual-relevant: 43 (19)<br>Auditory-relevant: 43 (20) | 25 (12) |
| Number of participants with a neurodevelopmental or psychiatric condition | 77 autism | 31 total<br>5 = ADHD<br>2 = anxiety disorder<br>20 = autism<br>4 = BAP | 33 total<br>7□=□ADHD<br>2□=□anxiety disorder<br>20□=□autism<br>3□=□BAP<br>1□=□bipolar | - | - |
| Age in years, mean (st. dev.) | 10.45 (1.8) | 11.7 (2.8) | 11.59 (2.87) | Visual-relevant: 22.40 (4.18)<br>Auditory-relevant: 22.28 (3.66) | 22.79 (3.54) |
| IQ | 113.7 (15.1) | 107.8 (15.1) | 107.23 (15.93) | - | - |
| fMRI data type | Resting-state | Continuous attention task (gradCPT) | Continuous attention task (gradCPT) | Continuous attention task (avCPT) | Continuous attention task (gradCPT) |
| Functional acquisition sequence features | Multisite study; scanner strength and acquisition parameters varied | 3 T Siemens Prisma; multiband acquisition sequence (acceleration factor = 5); TR = 1 s; voxel size = 2 mm<sup>3</sup> | 3 T Siemens Prisma; multiband acquisition sequence (acceleration factor = 5); TR = 1 s; voxel size = 2 mm<sup>3</sup> | 3T Philips Ingenia; multiband acquisition sequence); TR = 1 s; voxel size = 2.526 mm × 2.526 mm × 3 mm | 3 T Siemens Trio TIM; multiband acquisition sequence (acceleration factor = 2); TR = 1 s; voxel size = 2.5 mm<sup>3</sup> |
| Preprocessing features and head motion approach | BioImage Suite (Joshi et al. 2011), SPM8 motion correction; 24-parameter motion model (Satterthwaite et al. 2013); global signal regression | BioImage Suite (Joshi et al. 2011), SPM8 motion correction; 24-parameter motion model (Satterthwaite et al. 2013); global signal regression | BioImage Suite (Joshi et al. 2011), SPM8 motion correction; 24-parameter motion model (Satterthwaite et al. 2013); global signal regression | AFNI (Cox 1996); 24-parameter motion model (Satterthwaite et al. 2013); global signal regression | BioImage Suite (Joshi et al. 2011), SPM8 motion correction; 24-parameter motion model (Satterthwaite et al. 2013); global signal regression |
| Original connectome details | Shen 268-node functional atlas (Shen et al. 2013); 35,778 unique edges | Shen 268-node functional atlas (Shen et al. 2013); 35,778 unique edges | Shen 268-node functional atlas (Shen et al. 2013); 35,778 unique edges | Shen 268-node functional atlas (Shen et al. 2013); 35,778 unique edges | Shen 268-node functional atlas (Shen et al. 2013); 35,778 unique edges |
| Amount of fMRI data | One four-minute resting-state run | Two 4 min gradCPT runs averaged | Two 5:02 gradCPT runs averaged | One ten-minute visual-relevant CPT, one ten-minute auditory-relevant CPT | Three 13-minute 44-second gradCPT runs concatenated |
| Phenotype predicted in original model | SRS total scores (Constantino et al. 2003) | ADOS total calibrated score (Lord et al. 2012) | Performance on attention task (*d*’) | Performance on attention task (*d*’) | Performance on attention task (*d*’) |
| Number of trials in CPT | - | 480 | 404 | 500 | 2700 |
| Prediction performance in original sample | Pearson’s r between predicted and observed total SRS scores = 0.32, P-value < 5E-10 | Spearman’s rho between predicted and observed total ADOS scores = 0.445, P-value = 0.002 | Spearman’s rho between predicted and observed total d’ scores = 0.54, P-value = 0.0001 | - | Pearson’s r between predicted and observed total d’ scores = 0.84, P-value = 1.3E-7 |
| Phenotypes successfully predicted in test datasets in original publication | ADHD Rating Scale IV (DuPaul 1998) | Sustained attention task performance (d’) (Rosenberg et al. 2016)<br>SRS scores (Constantino et al. 2003) | Sustained attention task performance (d’) (Rosenberg et al. 2016) | - | ADHD Rating Scale IV (DuPaul 1998) |

Regarding aim one, we hypothesized that we would detect overlap across all of the attention and autism models, because models trained to predict autism symptoms contain information about sustained attention (and vice-versa). That is, variance in autistic traits tends to correlate with sustained attention (e.g., Corbett and Constantine 2006; Lin et al. 2023; Lake et al. 2019; Horien et al. 2023; Horien et al. 2026). In addition, all of the models in the present paper relate to the ability to sustain attention in some way (Figure 1), suggesting that similar brain networks might be involved. Regarding aim two, we hypothesized we would observe neurobiological commonalities within-phenotype.

## Methods

### Overview of the methodological approach

In this paper, we used data from five previously published studies (‘publication preregistration’; Rosenberg et al. 2020) that have built predictive models using CPM (Figure 1). Despite differences in the datasets, all models were related to sustained attention, including those derived from samples comprising participants with autism. However, it should be kept in mind that the exact relation to attention differed across samples (see Figure 1E for the relation to attention, as well as the exact phenotype predicted by each model). A brief description of the original dataset used to generate the model is offered below; see Supplemental Methods and Table 1 for full details.

The five models were selected because they contained the following similarities allowing comparisons across studies. First, the same functional brain atlas was used (Shen et al. 2013). Second, all studies used connectome-based predictive modelling (CPM) (Finn et al. 2015; Shen et al. 2017), a machine learning method for deriving brain-behavior models from functional connectomes. To establish the upper bounds of consistency, we leveraged other similarities. Multiple models were derived using similar attention task scanning conditions. Some of the models were validated on the same external samples (Supplemental Figure 1). Further, two models from separate publications shared approximately 85% of subjects, but predicted different phenotypes—one predicting attention task performance and the other predicting autism symptoms.

### Description of datasets used to generate predictive models

All datasets were collected in accordance with the institutional review board (IRB) or research ethics committee at each site and all studies were approved by the IRB at each site (Yale University IRB IORG number IORG0000431; University of Chicago IRB IORG number IORG0000201). Where appropriate, informed consent was obtained from the parents or guardians of participants. Written assent was obtained from children aged 13–17 years; verbal assent was obtained from participants under the age of 13 years.

#### Dataset 1, model 1: ABIDE model

The Autism Brain Imaging Data Exchange (ABIDE) (Di Martino et al. 2014; Di Martino et al. 2017) dataset comprised individuals with and without autism (Table 1). Throughout the text, we refer to this sample as the ‘ABIDE model/sample’. Full exclusion criteria have been described previously (Lake et al. 2019).

#### Dataset 2, model 2: Yale youth autism model

The second dataset comprised 63 participants (Table 1); full exclusion criteria are available elsewhere (Horien et al. 2026). Note that in addition to autism diagnoses in this sample, four participants were classified as belonging to the broader autism phenotype (Ingersoll 2010). Hereafter, we refer to this model/sample as the ‘Yale youth autism model/sample.’

#### Dataset 2, model 3: youth attention model

The second dataset was also used to generate a different model. The sample comprised 70 subjects (Table 1); full details of the sample have been described elsewhere (Horien et al. 2020; Horien et al. 2023). We refer to this model/sample as the ‘youth attention model/sample,’ but readers should keep in mind the transdiagnostic nature of the dataset. Note that because they were drawn from the same parent studies, models 2 and 3 shared 56 participants.

#### Dataset 3, model 4: avCPT model

The next dataset has been described elsewhere (Corriveau et al. 2025). The final sample size was n = 43 (Table 1). Participants completed an in-scanner 10-minute audio-visual continuous performance task (avCPT; we refer to this model/sample hereafter as the ‘avCPT model/sample’). We used the visual condition in the main analyses; data from the auditory condition were used during control analyses when performing representational similarity analysis (please see “Assessing brain-behavior relationships using representational similarity analysis” below).

#### Dataset 4, model 5: adult attention model

The last dataset comprised n = 25 neurotypical adults (hereafter referred to as the ‘adult attention model/sample’ (Table 1); full details of this sample are described elsewhere (Rosenberg et al. 2016).

### Functional imaging conditions: sustained attention tasks and resting-state

Functional imaging data were obtained while participants in the Yale youth autism sample and the youth attention sample completed the gradCPT in the scanner (Esterman et al. 2013; Rosenberg et al. 2013; Rosenberg et al. 2016). Briefly, participants viewed grayscale pictures of cities and mountains, with images transitioning from one to the next every 1,000 ms. Subjects were instructed to respond with a button press for city scenes and to withhold button presses for mountain scenes. City scenes occurred randomly 90% of the time. Performance was calculated using *d’* (sensitivity), the participant’s z-scored hit rate minus z-scored false alarm rate.

Subjects in the adult attention sample (Rosenberg et al. 2016) also performed the gradCPT in the scanner. The same parameters were used as above, except scene transitions took 800 ms. Participants in the avCPT sample completed an audio-visual continuous performance task (avCPT) (Corriveau et al. 2025), with a separate auditory and then a visual run(see Supplemental Methods for full details). In ABIDE, functional data comprised resting-state scans as described previously (Di Martino *et al*. 2014; Di Martino *et al*. 2017).

### Processing of functional imaging data

Functional data were processed as previously described (Greene et al. 2018; Horien et al. 2018; Rapuano et al. 2020; Greene et al. 2022; Corriveau *et al*. 2025) and followed the same general approach. This included alignment of functional data to a shared space, regression of covariates of no interest, including a head motion model, the mean signal from white matter and ventricle masks, and mean global signal (see Supplemental Methods, ‘Preprocessing of imaging data’ for full details).

Connectomes were generated using a 268-node atlas (Shen et al. 2013). The mean time-course of each region of interest (“node”) was calculated, and the Pearson correlation coefficient was calculated between each node pair to achieve a symmetric 268×268 matrix of correlation values representing connections between nodes (“edges”). Nodes were grouped into one of ten canonical functional networks described previously (Noble et al. 2017; Horien et al. 2019). Care was taken in each study to ensure artifacts related to head motion were not driving results (see Supplemental Methods, ‘Minimizing the impact of head motion’ and Supplemental Table 1).

In the avCPT sample, some subjects were missing edges due to incomplete coverage during the functional scan. If any subject was missing an edge, we removed that edge from all participants being compared from all samples; the final connectome size was 32,640 edges and 31,626 edges in the auditory and visual runs, respectively.

### Connectome-based predictive modelling

Connectome-based predictive modelling (CPM; Supplemental Figure 2) was used to generate the models in the current work. Importantly, each original publication included controls to ensure that variables such as age, sex, head-motion, and other factors were not confounding CPM results (Rosenberg et al. 2016; Lake et al. 2019; Horien et al. 2023; Corriveau et al. 2025; Horien et al. 2025; Horien et al. 2026). CPM uses cross-validation to determine the functional connections predictive of a given phenotype, resulting in a summary network model. Based on the edges most positively and negatively correlated with behavior, positive and negative network models are generated, respectively. The summary network model is then used to predict phenotypic outcomes in unseen participants. The summary network model is the output of interest in the present work. Due to the relationship between autism and attention (higher autism symptoms are associated with poorer attention (Allen and Courchesne 2001; Landry and Parker 2013; Lyall et al. 2017), we considered the networks from ABIDE and the Yale youth autism study in the opposite direction. That is, the positive ABIDE and Yale youth autism networks were considered with the negative networks from the avCPT, adult attention, and youth attention samples, and vice-versa.

### Accounting for differences in the size of the previously published predictive models

Because of differences in modelling parameters in each original study (Supplemental Table 2), each summary network in the original publication was a different size. To ensure that this did not confound results, we re-analyzed the original data used to construct the models in each sample. Edges were correlated with behavior in each sample, ranked according to correlation strength, and the top 1,000 positively and negatively correlated edges were retained for analysis. (This corresponds to the feature selection step in Supplemental Figure 2, box 3, though instead of a feature selection threshold, we retained only the top 1,000 edges.) Note that 2,000 edges are chosen in total, in line with the original networks studied here, as well as previously published CPM networks of other phenotypes (e.g., Cheng et al. 2025). To ensure results were not driven by the use of a specific threshold, we also tested 500 edges in each positive and negative network (1,000 total), as well as 2,500 edges in each network (5,000 edges total).

### Statistical overlap of edges in predictive models

To assess the probability of networks containing a statistically significant number of shared edges, we performed 1,000 iterations of generating random networks, ensuring the null networks were the same size as the original network from a given sample (see Supplemental Figure 3A for a schematic of the process). We then compared the number of times the random networks resulted in a number of overlapping edges greater than or equal to the observed number of overlapping edges in the actual predictive networks. In other words, we calculated the overlap of random network models and determined how many edges were shared by models.

We next assessed the overlap of edges among specific models themselves in a pairwise fashion. That is, we considered two predictive models at a time and asked if the overlap among edges was statistically significant. To do so, we used the hypergeometric cumulative density function (CDF) as previously described (Rosenberg et al. 2020; Corriveau et al. 2025). We used the Benjamini-Hochberg procedure (Benjamini and Hochberg 1995) to control for multiple comparisons (20 total, ten comparisons among the positive networks and ten among the negative networks). (See Supplemental Methods, ‘Statistical overlap of edges in predictive models: CDF’ for a full description of the procedure.)

### Quantifying the contribution of network pairs to predictive models

To determine the importance of specific networks in the predictive models, we quantified the number of edges between a given network pair. We assessed the top 1,000 edges correlated positively and the top 1,000 edges correlated negatively with behavior. Specifically, we grouped the nodes linking each edge into one of ten networks (Noble et al. 2017), resulting in a 10×10 symmetric matrix (focusing our analyses on the diagonal and bottom half of the matrix, due to the symmetry). The number of edges linking into a given network pair was then quantified, resulting in an integer in each cell of the matrix. The integer was divided by the total number of edges in the network pair, to account for network size as previously reported (Horien et al. 2019; Horien et al. 2023). For each individual model, we assessed statistical significance by calculating null predictive network models for each sample (see Supplemental Figure 3B for a schematic of the process). As each individual model involved 55 comparisons per positive network and 55 comparisons per negative network, we used the stricter Bonferroni correction (Bland and Altman 1995) when reporting significant *P*-values for a network pair (correcting for 55 tests). Having determined which network pairs were statistically significant at the individual model level, we then assessed which networks were statistically significant in all five datasets (Figure 6B), in those predicting autistic traits (Figure 6C), and in those predicting continuous attention task performance (Figure 6D). To ensure results were robust, we repeated analyses using 500 edges in each positive and negative network (1,000 total), as well as 2,500 edges in each network (5,000 edges total).

### Assessing brain-behavior relationships using representational similarity analysis

To assess the importance of different dataset features on the brain-behavior relationships, we used representational similarity analysis (RSA) (Kriegeskorte et al. 2008). In each sample, we correlated each edge in the connectivity matrices from all participants with the behavior of interest, resulting in a 31,626×1 vector per dataset. (Note that the visual run of the avCPT sample was used for our main analysis in RSA.) After correlating these vectors across samples to obtain a feature importance similarity matrix, we generated model representational similarity matrices based on mean age of the sample, functional run type (task or rest), behavioral measure (attention score or autism symptom score), and percentage of the sample diagnosed with autism. We *z*-scored the entries in each representational similarity matrix to obtain standardized betas in a multiple linear regression, allowing us to assess the unique variance in the feature importance similarity matrix explained by each predictor:

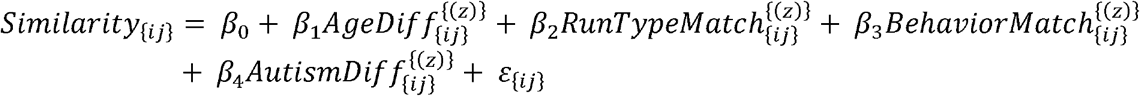

where *Similarity*_{*ij*}_ is the Pearson correlation between feature importance vectors for datasets *i* and *j*; Age *Dif f*_{*ij*}_ is the absolute difference in mean age between datasets; *RuntypeMatch_{ij}_* is a binary indicator of the type of functional data used, either resting-state or task data (1 = same run type, 0 = different);*BehaviorMatch_{ij}_* is a binary indicator of the type of phenotype predicted, either autistic symptoms or attention task performance (1 = same behavioral measure, 0 = different behavioral measure); *AutismDif f*_{*ij*}_ is the absolute difference in percentage of participants diagnosed with autism between datasets; *β*_0_ represents the intercept; and *ε*_{*ij*}_ is the residual error. To ensure results were robust, we repeated analyses using the auditory run of the avCPT sample.

### Code/data availability and generative AI statement

Preprocessing of the original datasets (as well as glass brain and circle plot visualizations) used freely available software: https://medicine.yale.edu/bioimaging/suite/. CPM code used to generate the original predictive models is available here: https://github.com/YaleMRRC/CPM. Template scripts used for preprocessing and analysis of the functional data are available here, as are the node-to-network labels: https://github.com/clhorien/transdiagnostic_attn_overlap/tree/main. The functional parcellation is available here: https://www.nitrc.org/frs/?group_id=51. ABIDE data are available here: https://fcon_1000.projects.nitrc.org/indi/abide/. Data from the avCPT sample are available here: https://osf.io/bt2xy/overview. All other data or materials are available from the authors upon request. A generative AI/large language model (LLM) was not used to write this paper.

## Results

### Characteristics of previously published predictive models

We began by assessing basic aspects of the previously published predictive models. Specifically, we analyzed characteristics of the CPM networks derived from each study (i.e., we did not require networks to be the same size at this stage). The original networks comprised a collection of edges spanning the entire brain (Supplemental Figures 4-6). Network size and the number of interhemispheric versus intrahemispheric connections were also assessed. Despite using different modelling parameters (Supplemental Table 2), networks were generally the same size and comprised a similar number of connections within and between hemispheres (Supplemental Figure 4; Supplemental Table 3). Functional networks from across the brain contributed to the models, from visual, somatomotor, and cerebellar networks to higher-order association networks, including the default mode, frontoparietal, and medial frontal networks (Supplemental Figures 5-6). We did not observe qualitative differences between models built to predict autism traits compared to those built to predict sustained attention phenotypes.

### Edge overlap across all predictive models

We next quantified the number of times individual edges were shared across models. Because of differences in network size across samples, network size was kept consistent across samples. That is, instead of using the summary models from each publication, we determined the top 1,000 edges most strongly correlated with behavior in each direction (the ‘positive’ and ‘negative’ networks, respectively; recall that we accounted for autism diagnosis in this procedure). When we localized the edges, areas from across the brain were involved in the models (Figure 2). As we saw with the data shown in Supplemental Figures 5-6 and Supplemental Table 3, we observed that all lobes tended to be represented, with both right and left hemispheres, as well as intra- and inter-hemispheric connections present in the models. As before, there appeared to be no qualitative differences between autism-specific models (M1 and M2) compared to attention-specific models (M3 – M5).

**Figure 2.**
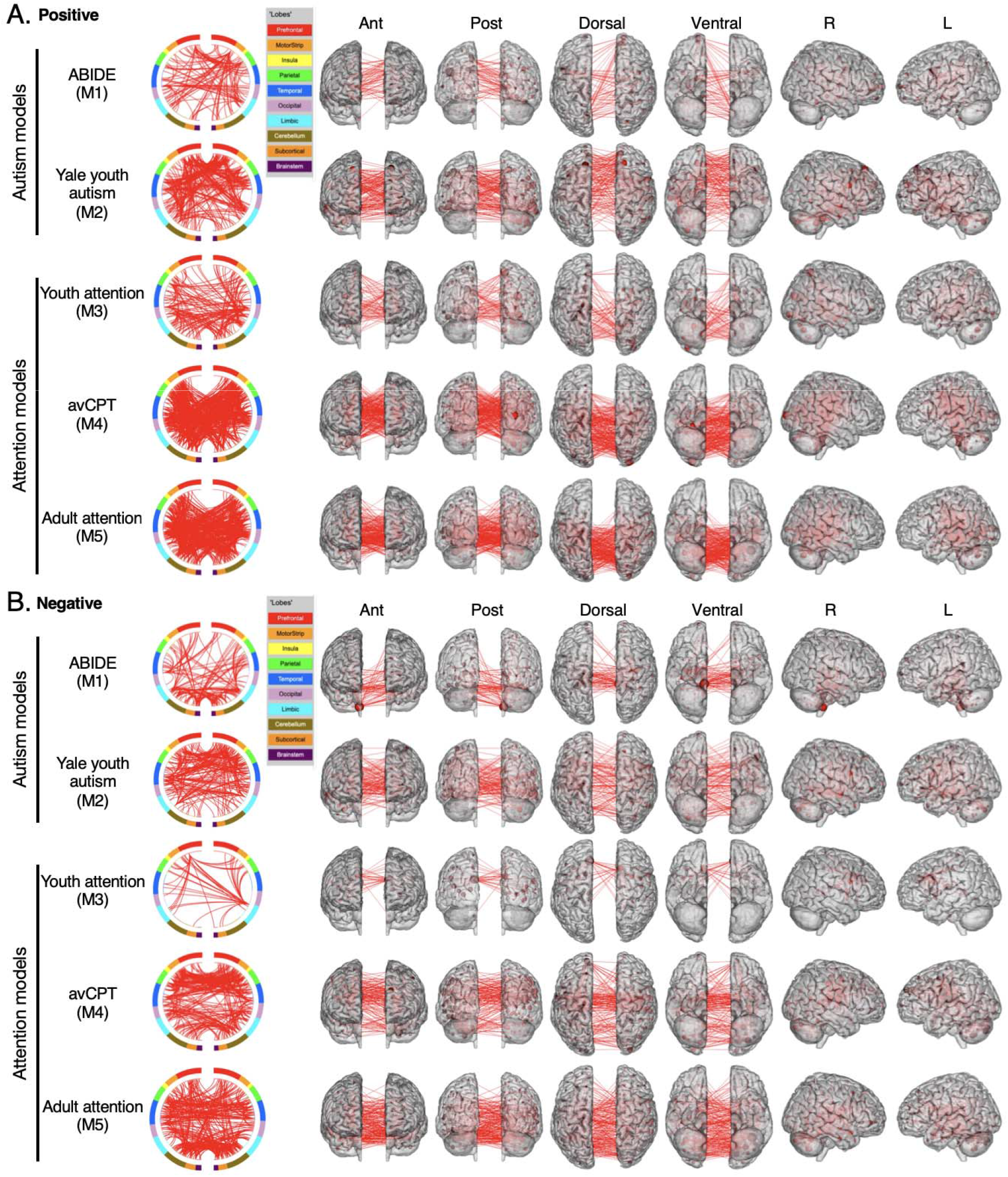
The network masks when assessing the top 1,000 edges correlated with behavior. A) Positive network masks. B) Negative network masks. Both panels: Each row corresponds to a different network model; the dataset from which the network was obtained is indicated to the left. A circle plot is shown immediately to the right of the dataset name. The topmost aspect of the circle represents anterior; the bottom, posterior. The right half of the circle plot corresponds to the right hemisphere of the brain. A legend indicating the approximate anatomic “lobe” is shown to the top right of the circle plots. The same edges are plotted in the glass brains as lines connecting different nodes; in these visualizations, nodes are sized according to degree, the number of edges connected to that node. The view of the brain is indicated at the top of the panel. To aid in visualization, we have thresholded the data to only show nodes with a degree threshold > 20. ABIDE, autism brain imaging data exchange; Ant, anterior; avCPT, audio-visual continuous performance task; L, left; M, model; Post, posterior; R, right. Alt text: A figure showing where edges are located on the brains for each dataset in the positive and negative network masks.

After creating new summary models, overlap was calculated across the models. The goal was to determine the probability that an edge was shared by *any* of the models. Hence, the model label was not used in this initial analysis. In the positive and negative networks, we observed that 408 and 423 edges were shared by two models, respectively (Figure 3A). Both of these values were statistically significant compared to the overlap found in null models (*P*-value for positive network = 0.00099; *P*-value for negative network = 0.00099, corrected for multiple comparisons). When assessing whether edges were shared across a greater number of models, 28 edges were shared across three models in the positive networks; 35 edges were shared across three models in the negative networks (*P*-value for positive network = 0.00099; *P*-value for negative network = 0.00099, corrected for multiple comparisons). Notably, no edges were shared across four models in the positive networks; only one edge was shared across four models in the negative networks, which was not statistically significant (*P*-value = 0.167, corrected for multiple comparisons). No edges were shared across five models in either network. To ensure results were consistent, we repeated analyses using a different number of edges. We observed a similar pattern when testing 500 edges in each network (1,000 total) and 2,500 edges in each network (5,000 total; Supplemental Figure 7).

**Figure 3.**
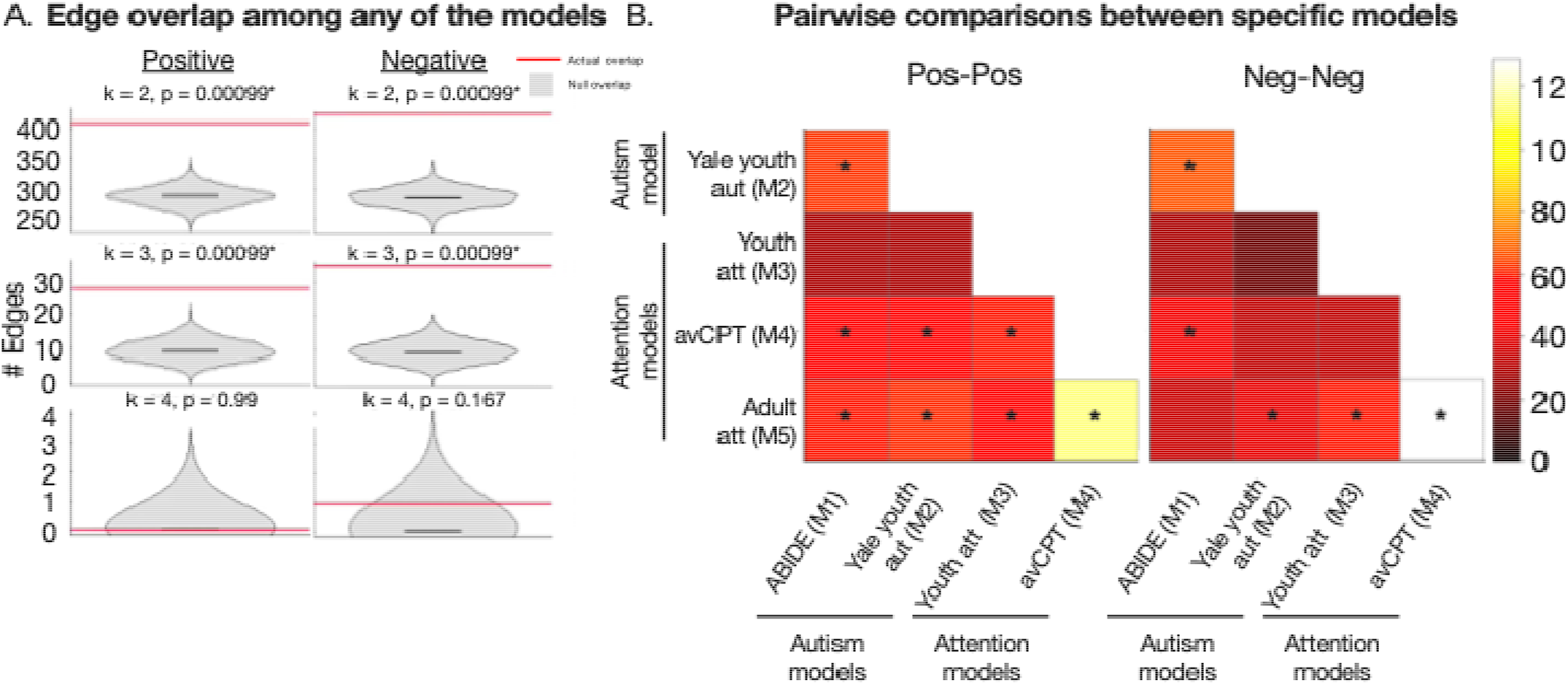
Quantifying the occurrence of individual edges across models. A) Violin plots show the number of edges shared across models as a red horizontal line in each plot. The grey violin plot represents null results of the number of shared edges obtained from 1,000 permutations, with the black bar in the violin plot indicating the median of the 1,000 permutations. The positive network is shown in the left column; the negative network, in the right column. Above each plot, ‘k’ indicates the number of models in which an edge occurred. *P*-values are shown at the top of each plot; asterisks (*) indicate statistical significance after multiple comparisons correction. Note that the aim in this panel is to establish the probability that an edge is shared by *any* of the models. In this analysis, the model label is not used. B) Pairwise comparisons between specific models. The model label is now used, as the analysis asks which models share a statistically significant number of edges. Two matrices are shown: overlap between the top-ranked edges (‘Pos-Pos’, left) and overlap between the bottom-ranked edges (‘Neg-Neg’, right). Each cell corresponds to the number of edges shared between models. The dataset from which the model was obtained is indicated along the rows and columns of each matrix. The colorbar is scaled so that fewer shared edges (in terms of absolute number) are darker colors; more shared edges are lighter colors. An asterisk indicates a statistically significant number of shared edges between the two models after correcting for multiple comparisons. ABIDE, autism brain imaging data exchange. Att, attention; Aut, autism; avCPT, audio-visual continuous performance task; Neg, negative network; Pos, positive network. Alt text: Graphs showing the degree of overlap among edges in any of the models using violin plots, as well as matrices showing pairwise comparisons between the different network masks.

### Edge overlap when considering the models pairwise

Having observed that edges were shared by some of the models, we next assessed the models pairwise. Note that the aim in this analysis was complementary to the aim shown for Figure 3A, in which the goal was to determine if edges were shared, regardless of model identity. We built on the observation that edges were shared by investigating all possible pairwise combinations of the models to more clearly delineate which models shared a statistically significant number of edges (Figure 3B).

In the positive network, we observed 8/10 pairwise comparisons shared a statistically significant number of edges (all significant comparisons shared >47 edges, all *P*-values < 0.003, corrected for multiple comparisons). The 2/10 pairwise comparisons that did not share a statistically significant number of edges both comprised models predicting incongruent phenotypes (i.e., when one model predicted autism traits and the other predicted attention). That is, the ABIDE (M1) model predicting an autistic trait (SRS) and the youth attention model (M3) predicting attention (gradCPT task performance) did not share edges (shared edges = 31, *P*-value = 0.57). The other non-significant comparison involved the samples sharing subjects, but predicting different phenotypes (i.e., the Yale youth autism model (M2) predicting ADOS and the youth attention model (M3) predicting gradCPT task performance; shared edges = 32, *P*- value = 0.49).

In the negative network, 5/10 pairwise comparisons shared a statistically significant number of edges (all significant comparisons shared >42 edges, all *P*-values < 0.019, corrected for multiple comparisons). Of these, 4/5 non-significant comparisons involved incongruent phenotypes (Figure 3B). Of note, the two models sharing subjects but predicting different (incongruent) phenotypes (the Yale youth autism model, M2 and the youth attention model, M3) again did not share a statistically significant number of edges (shared edges = 20, *P*-value = 0.98).

To ensure findings were robust, we repeated pairwise comparisons with different network sizes. We obtained a similar pattern when testing 500 edges in each network and 2,500 edges in each network (Supplemental Figure 8). Specifically, overlap between models was detected pairwise. As with the data in Figure 3B, statistically significant overlap seemed to be associated with phenotypic congruence (i.e., when both models predicted autism or sustained attention phenotypes), and models sharing a statistically non-significant number of edges tended to be from incongruent phenotypes (although the relationship was not perfect). Taken together, the analyses indicated that while we did not observe a core edge set across all five models, most models tended to have overlapping edges when considered pairwise and phenotypes were congruent.

### Neuroanatomical visualizations of shared edges

For neurobiological context, we localized shared edges across all five of the models when there were 1,000 edges in each network (Figure 4); nodes with high degree (indicating many edges were connected to that node) were localized as well (Figure 5). We also determined the location of the high degree nodes shared across the five models, their corresponding MNI coordinates, as well as the hemisphere and anatomic lobe to which each node belonged (Supplemental Table 4). Finally, we visualized edges that were shared when models predicted congruent phenotypes (Supplemental Figures 9 and 10). Specifically, we considered within-phenotype overlap for the edges that were shared for the autism models (ABIDE, M1; and Yale youth autism, M2), as well as the within-phenotype overlap for the edges that were shared for the attention models (Youth attention, M3; avCPT, M4; and adult attention, M5).

**Figure 4.**
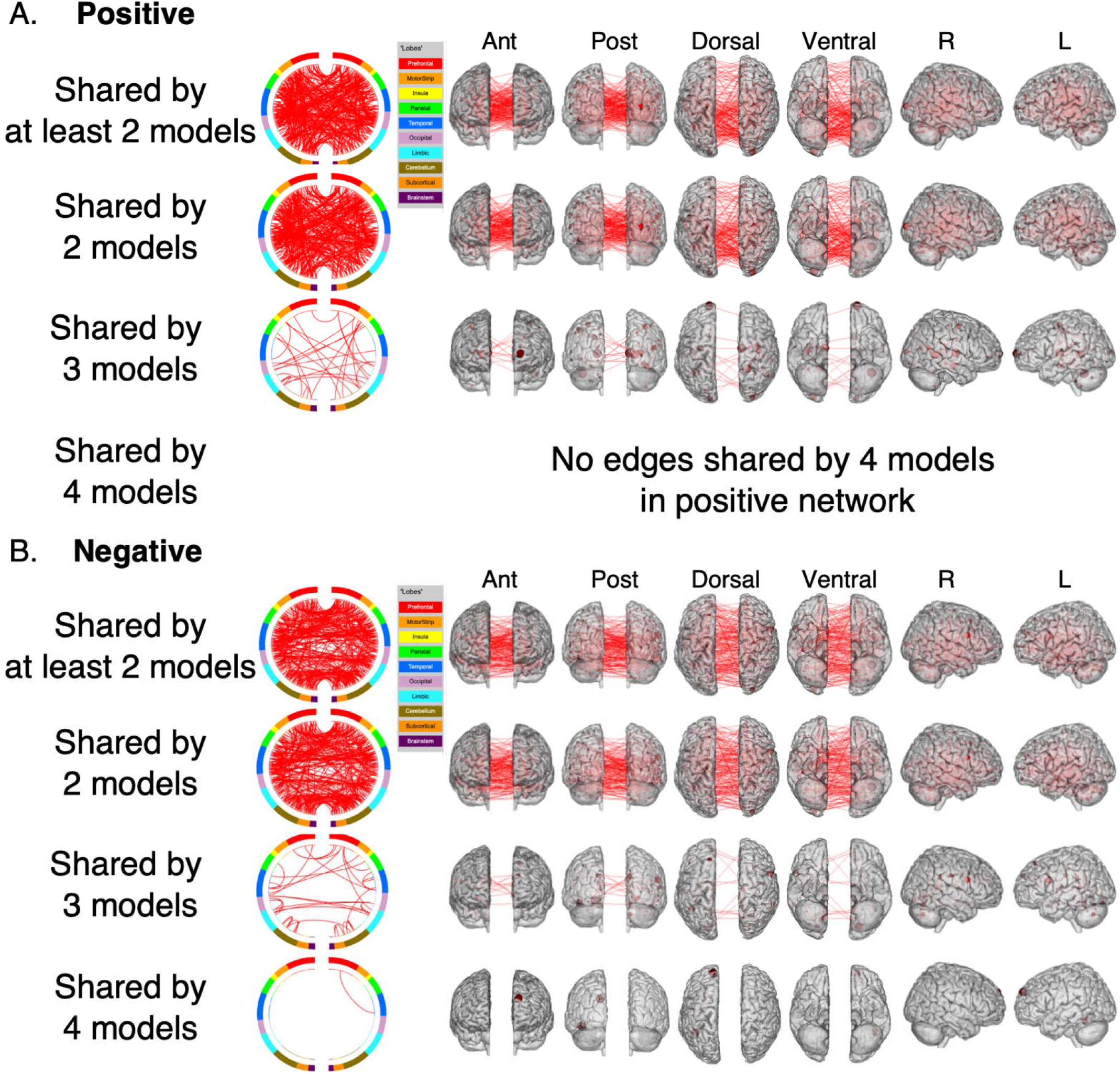
Determining the anatomical location of shared edges in the positive and negative network models. A) Positive network masks. B) Negative network masks. Shown in each row is the number of times an edge is shared. A circle plot is shown immediately to the right of the label indicating the number of times an edge is shared. The topmost aspect of the circle represents anterior; the bottom, posterior. The right half of the circle plot corresponds to the right hemisphere of the brain. A legend indicating the approximate anatomic “lobe” is shown to the top right of the circle plots. The same edges are plotted in the glass brains as lines connecting different nodes; in these visualizations, nodes are sized according to degree, the number of edges connected to that node. The view of the brain is indicated at the top of the panel. ABIDE, autism brain imaging data exchange; Ant, anterior; avCPT, audio-visual continuous performance task; L, left; M, model; Post, posterior; R, right. Alt text: Circle plot and glass brain representations showing where edges were shared in the positive and negative networks, with edges shown on the brain, as well as which nodes they are connected to.

**Figure 5.**
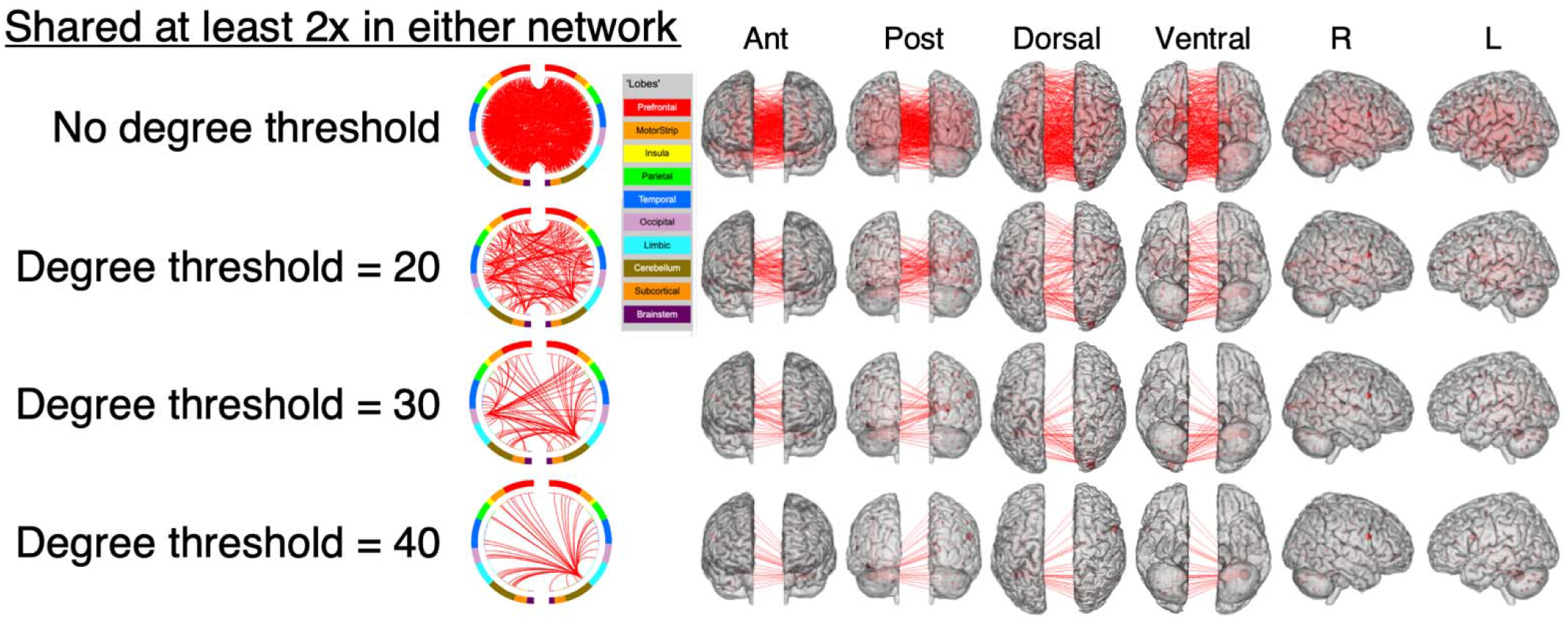
Determining the anatomical location of shared edges of both network masks. Here, we are plotting an edge from both the positive and negative networks if it is shared at least twice in either of the networks. The top row indicates the visualization is shown without applying a degree threshold. The rows beneath represent the same data, with different degree thresholds applied (with degree corresponding to the number of edges connected to that node). A circle plot is shown immediately to the right of the label indicating the degree threshold. The topmost aspect of the circle represents anterior; the bottom, posterior. The right half of the circle plot corresponds to the right hemisphere of the brain. A legend indicating the approximate anatomic “lobe” is shown to the top right of the circle plots. The same edges are plotted in the glass brains as lines connecting different nodes; in these visualizations, nodes are sized according to degree. The view of the brain is indicated at the top of the panel. ABIDE, autism brain imaging data exchange; Ant, anterior; avCPT, audio-visual continuous performance task; L, left; M, model; Post, posterior; R, right. Alt text: Circle plot and glass brain representations showing where edges were shared at least twice in the models. Results are thresholded according to degree, with a higher degree threshold resulting in a sparser number of edges.

**Figure 6.**
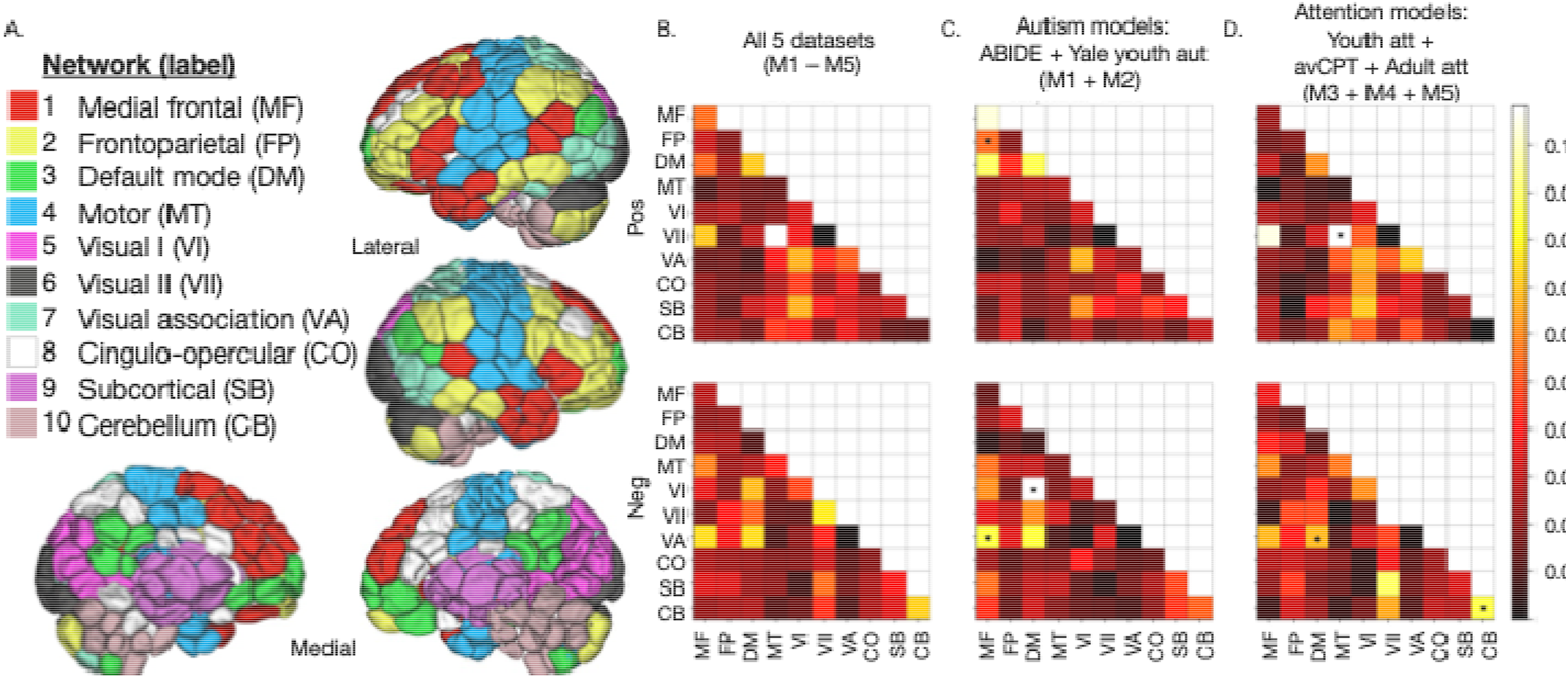
Neuroanatomy of the predictive network models. A) Network labels used in the current work. A 268-node functional atlas was used. Nodes are shown in lateral and medial views on brain images (right and lower portion of panel A). Nodes were grouped into 10 functional networks (left upper portion of panel A). Network label abbreviations are shown in parentheses after each network name; network abbreviations are used in panels B-D of the current figure. B) Summary matrix for all five datasets. C) Summary matrix for the datasets predicting autistic phenotypes. D) Summary matrix for the datasets predicting continuous attention task performance. For all plots: Positive network matrices are shown in the top row; negative network matrices, in the bottom row. Each cell in a matrix corresponds to the fraction of edges in a given network pair that were included in the given model, after correcting for network size. The colorbars for each matrix are scaled so that a lower fraction of edges corresponds to darker colors; a higher fraction of edges corresponds to lighter colors. Statistically significant network pairs after multiple comparisons correction are denoted with an asterisk (*). ABIDE, autism brain imaging data exchange; Att, attention; Aut, autism; avCPT, audio-visual continuous performance task; M, model; Neg, negative network; Pos, positive network. Network labels: MF, medial frontal; FP, frontoparietal; DM, default mode; MT, motor; VI, visual I; VII, visual II; VA, visual association; CO, cingulo-opercular; SB, subcortical; CB, cerebellum. Figure 6A adapted with permission from Horien et al. (2019). Alt text: A figure showing the resting-state functional networks comprising the models. Node-to-network labels are shown on the brain, along with matrices showing networks involved in all models, in the autism-specific models, and the sustained attention models.

Altogether, the data indicated that shared edges tended to be distributed widely across the brain, both when edges were shared across all models and within-phenotype. Models were spatially complex: there was no one lobe or region dominating the networks; models comprised both short- and long-range connections; and models comprised interhemispheric as well as intrahemispheric connections. Most functional networks contributed to the models, including sensorimotor networks and higher-order networks, such as the default mode, frontoparietal, and medial frontal networks. No clear hemispheric differences (right versus left) emerged. There were no apparent differences in the location of edges when the positive and negative networks were compared.

Interestingly, the cerebellum contained a large number of high degree nodes in both the positive and negative networks (Supplemental Table 4), with the same node in the left cerebellum containing the highest degree in both the positive and negative networks when considering edges shared by at least two models (Node 238 in Shen-268 parcellation; MNI coordinates = -36.95, -52.94, -31.11; Positive network: degree = 21; Negative network: degree = 22). In addition, when considering edges shared by at least two models, the positive and negative networks shared 4/15 (26.6%) of the top nodes, suggesting that a few key nodes might be responsible for the similarities that were detected between autism and sustained attention models in the present work. See Supplemental Table 4 for more details across all overlap thresholds, as well as Supplemental Figures 9 and 10 for within-phenotype overlap.

### Network consistency in brain-phenotype relationships

In the previous section, we asked if individual edges were themselves shared across models. In this section, we assessed the models at the network level. Such an approach was designed to address if a similar proportion of edges in a given network were involved across multiple models, regardless of whether the same edges were shared across models. We again assessed the top 1,000 edges that were positively correlated with behavior and the top 1,000 edges that were negatively correlated with behavior. We then determined to which functional networks the edges belonged. Specifically, we assessed the locations of edges within and between functional networks, asking whether the same network pairs were significantly overrepresented across models.

Because of differences in the sizes of within- and between-network pairs in the connectomes, we corrected for network size in this procedure (Methods). We found that network pairs (shown in Figure 6A) linking regions across the brain were represented (Figure 6B). In the positive network, visual, motor, and default mode networks had the highest fraction of edges across the five samples. In the negative network, visual, default mode, and cerebellar networks had the highest fraction of edges. We obtained similar results when testing network sizes of 500 edges in each network and 2,500 edges in each network (Supplemental Figure 11).

Within the individual samples, the location of statistically significant network pairs was assessed after accounting for multiple comparisons (see Supplemental Figure 3B for a schematic of the procedure). No discernible pattern emerged regarding the location of statistically significant network pairs in the individual samples (see Supplemental Figure 12 for the network pairs surviving multiple comparisons correction). Sensorimotor and cerebellar networks were statistically significant, as were higher-order networks, including the default mode and frontoparietal networks. Findings were similar when different-sized networks were used (500 and 2,500 edges; Supplemental Figure 12A-C).

There was little overlap in terms of which network pairs were statistically significant across the samples as a whole. For example, no network pairs were common across all five samples in the positive or negative networks (that is, none were consistently statistically significant; Figure 6B). When we considered overlap among the models predicting autistic phenotypes (Figure 6C), we observed three shared network pairs that were consistently statistically significant: one in the positive and two in the negative network. In the positive network, the shared network pairs involved medial frontal to frontoparietal networks. In the negative network, the shared network pairs involved medial frontal to visual association network and default mode to visual I network. There were three common network pairs when considering the models predicting continuous attention performance (Figure 6D). In the positive network, the shared network pair involved motor to visual II network. In the negative network, there were two shared network pairs: one involved default mode to visual association network and the other involved within-network connections in the cerebellum network.

We also assessed pairwise comparisons of shared network pairs and determined most pairs of samples shared at least one statistically significant network pair. Results were again consistent using the differently sized networks (Supplemental Figure 12D-F). The samples sharing subjects but predicting different phenotypes (Yale youth autism and youth attention) tended not to share network pairs, again suggesting that phenotypic incongruence was associated with different functional network involvement.

To confirm specificity of the positive and negative networks, as well as the relevance of phenotypic congruence, we flipped the sign of each network and assessed overlap. That is, the positive networks of the autism samples (ABIDE, M1; and Yale youth autism, M2) were compared with the negative networks of the attention samples (youth attention, M3; avCPT, M4; and adult attention, M5) and vice-versa. We observed no statistically significant shared network pairs in either case.

The results in this section suggested that the brain-phenotype models had distributed network representations. Although no core set of networks was observed across autism and attention models, some degree of overlap was detected when we considered congruent phenotypes and when we considered the datasets in a pairwise fashion.

### Exploring the factors associated with brain-phenotype relationships

Finally, we conducted a post-hoc investigation into which factors were associated with similarities among the models. In the previous sections, we found that although overlap was not absolute, some of the models shared features, especially when the predicted phenotypes were congruent. A natural follow-up question was whether there were meaningful differences between the datasets—such as demographic and other dataset factors—that were associated with model similarity. Therefore, we conducted analyses assessing brain-phenotype relationships using unthresholded data and RSA.

One advantage of RSA (Kriegeskorte et al. 2008) is the investigation of whole-brain patterns associated with a behavioral outcome, even those below the surface of what survives significance thresholding used in feature selection. We assessed predictors including the mean age of the sample, functional run type (task or rest), behavioral measure (attention score or autism symptom score), and percentage of participants in the sample with an autism diagnosis. Interestingly, we observed that there were no statistically significant relationships between the predictors and brain-phenotype similarity vectors (*P*-values > 0.05; see Table 2 for regression coefficients and confidence intervals).

**Table 2.** Results of RSA using unthresholded edge–behavior associations from each study. Shown are regression coefficients from models using z-scored predictors, along with the 95% confidence intervals (CI).

|  | Regression coefficient | Lower bound 95% CI | Upper bound 95% CI |
|---|---|---|---|
| Age | 0.55 | -1.61 | 2.71 |
| Run type | 0.026 | -0.085 | 0.14 |
| Behavioral measure | 0.069 | -0.21 | 0.35 |
| Autism diagnosis | -0.60 | -2.83 | 1.63 |

We also examined pairwise correlations among the datasets. The first step in RSA was to calculate a similarity matrix of edge-behavior vectors across datasets through the use of correlation. We compared the similarity among the samples to that observed from null models. Using the visual run of the avCPT task (M4), 4/10 pairwise dataset comparisons showed positive correlations (Figure 7; Supplemental Table 5; all Pearson correlations > 0.13; all *P*-values < 0.005, corrected for multiple comparisons). All of the positive correlations were between samples that had congruent phenotypes. One pairwise comparison showed a statistically significant negative correlation. This was between models with incongruent phenotypes (Yale youth autism sample (M2) and adult attention sample (M5); Pearson correlation = -0.14; *P*-value = 0.005, corrected for multiple comparisons; Figure 7).

**Figure 7.**
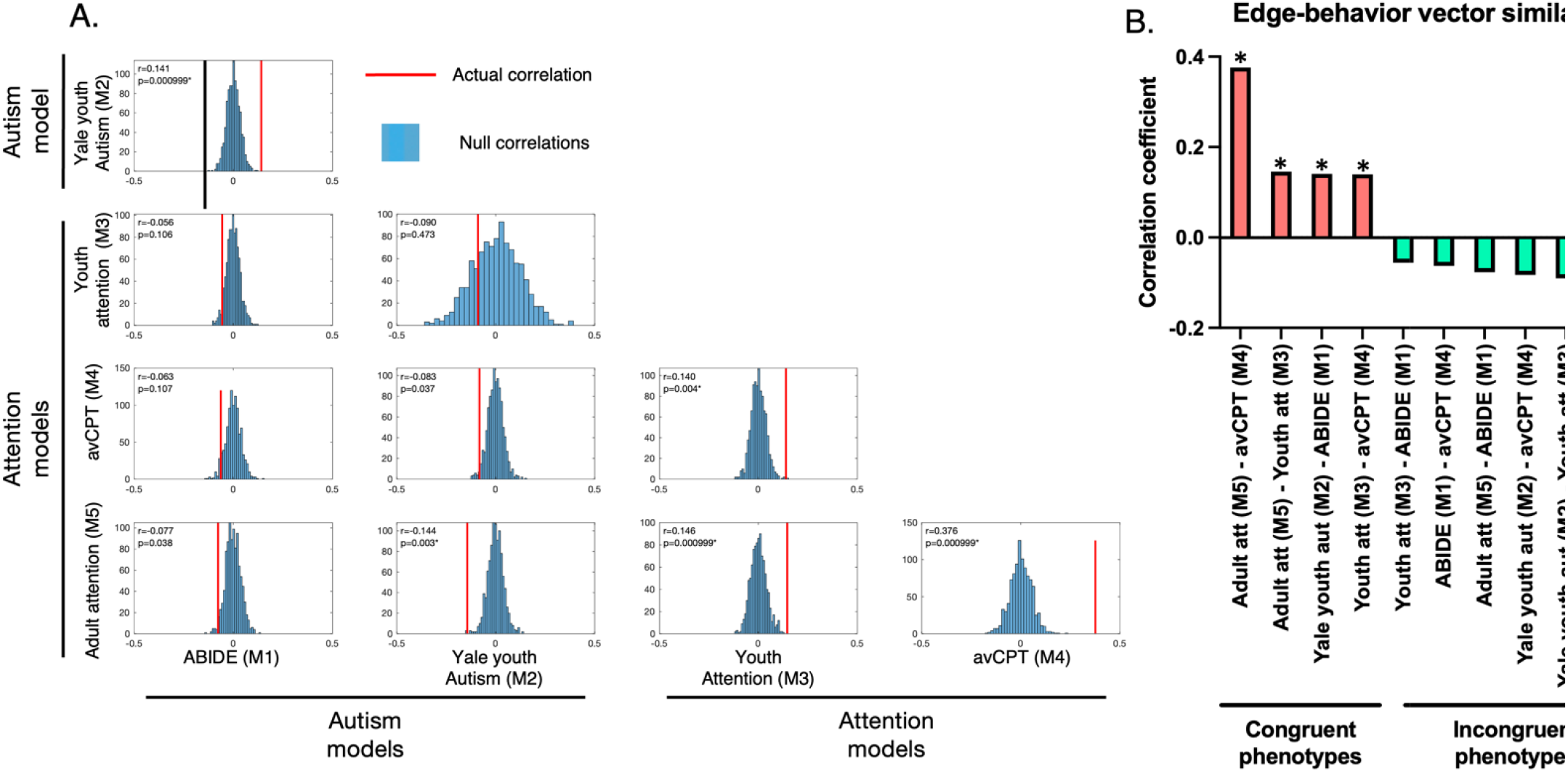
Relationship of brain-phenotype vectors across samples. A) Histograms showing the observed correlation (in red) between the brain-phenotype vectors (i.e., the Pearson correlation of the brain-phenotype vectors). Null values for the histograms are shown in blue. The datasets compared are indicated in the rows and columns beside the histograms. In the upper left of each plot, the Pearson correlation coefficient between the datasets in each row and column is shown, along with the *P*-value. B) Similarity of edge-behavior vectors across datasets. Each bar represents the Pearson correlation coefficient (y-axis) between the datasets indicated on the x-axis. Behaviors that are both related to attention or autism (‘Congruent phenotypes’) are indicated in salmon color; behaviors that are not related between datasets (‘Incongruent phenotype’), in which one sample comprises an autism phenotype with the other comprising an attention phenotype, are indicated in green. In both plots, an asterisk (*) indicates statistical significance after correction for multiple comparisons. ABIDE, autism brain imaging data exchange dataset; avCPT, audio-visual continuous performance task dataset. Alt text: A figure showing histograms describing the relationship between edge-behavior vectors, with actual data plotted against permuted data. A bar graph is shown summarizing the pairwise relationships, with congruent phenotypes (edge-behavior models where the phenotype is the same) showing positive correlations, whereas incongruent phenotypes (edge-behavior models where the phenotype is different) show negative correlations.

To ensure results were robust, we also tested similarity of the edge-phenotype vectors when the auditory condition from the avCPT sample (M4) was used instead of the visual condition (recall the avCPT sample comprised two separate conditions, one visual condition and one auditory condition). Findings were consistent (Supplemental Table 5). In sum, the factors influencing brain–behavior relationships in the present transdiagnostic samples were complex, although preliminary analyses (e.g., Figure 7B) suggested that models predicting similar phenotypes might share more similar features.

## Discussion

In the present work, we attempted to identify consistent functional connectome-based features associated with transdiagnostic models of sustained attention and autism traits. Using data from five previously published functional connectivity-based brain-behavior predictive models, we did not derive a core network. Nevertheless, we found that models did contain similar functional connections and resting-state functional networks when considered pairwise for congruent phenotypes. That is, models predicting sustained attention were more closely related to each other than models predicting autism symptoms and vice-versa. Preliminary results using RSA did not reveal a statistically significant relationship between brain-behavior vectors and factors including age and autism diagnosis. In sum, the data suggest that patterns predicting individual differences in autism and sustained attention tend to share some features, but the brain-behavior models of each appear to be phenotype specific.

### The lack of shared functional features across transdiagnostic models of attention and autism reinforces the importance of phenotype specificity

We did not identify a core predictive network across the five samples. The result is notable given the many similarities among the datasets. It is also notable because the models have generalized in similar (or in many cases the same) datasets. Furthermore, two of the models sharing approximately 85% of subjects, but predicting different phenotypes (ADOS scores in the Yale youth autism model, M2, and performance on the continuous attention task in the youth attention model, M3), did not contain overlapping edges or networks. One interpretation of the lack of overlap is that despite being broadly related to sustained attention, models tend to be specific to the construct they were built to predict. In other words, autism models tend to look more like autism models than sustained attention models, and vice-versa. This is in line with previous work investigating overlap in brain regions between autism and those who have difficulties with attention (e.g., ADHD), in that more similar brain areas are shared within diagnosis than across diagnoses (Christakou et al. 2013; Di Martino et al. 2013; Kernbach et al. 2018; Lukito et al. 2020; Tamon et al. 2024). Despite all of the opportunities available to find similarities among sustained attention and autistic models in the current work (using the same functional task in 4/5 studies, the same behavioral construct of *d*’ in 3/5 studies, and partially overlapping subject pools in the Yale youth autism and youth attention samples), we did not find commonalities, supporting the idea of phenotypic specificity.

The lack of strict overlap across all models might also relate to the spatially distributed nature of the models. The models comprised a sparse collection of edges spanning the entire brain, with inputs involving nearly every within- and between-network pair across subcortical, cortical, and cerebellar networks. Due to the sparsity, and the widespread nature of how the limited number of edges is distributed, overlap becomes more difficult. The multivariate nature of the models, made up of individual edges, also echoes results in the functional connectivity reliability literature. In particular, individual edges do not tend to have high reliability (Noble et al. 2017), but multivariate measures of reliability tend to be higher (Noble et al. 2019; Marek et al. 2022). Any individual edge might have a low chance of being shared across all of the models studied here due to low reliability.

### What are other potential explanations for the lack of shared features across all models?

Given the lack of similarity across the five models, it is reasonable to ask questions about other causes besides phenotypic specificity. For example, one could question the utility of CPM or whether it is useful at all to search for a core architecture in transdiagnostic autism and attention phenotypes. Regarding CPM, the approach involves selecting edges associated with a phenotype of interest, summarizing edge weights with a predictive model, and testing the model on unseen subjects (Finn et al. 2015; Shen et al. 2017). Similar to the CPM models described here, other successful CPM models generalizing out of sample tend to be sparse (containing <10% of the unique edges in a connectome) (e.g., Beaty et al. 2018; Rosenberg and Chun 2020; Feng et al. 2024). By definition, the feature selection step involves selecting edges most correlated with a phenotype. This implies phenotypic specificity to the edges that are selected: no argument is made about utility or generalizability to other phenotypes.

Assessing aspects such as biological utility or relevance to other phenotypes are post-hoc questions that can help shed light on cognitive processes, as well as their relevance to clinical conditions. For example, other groups have searched for overlap in CPM predictive network models and observed commonalities. In a study of approximately 1,100 teenagers and young adults (Cheng et al. 2025), the authors found overlap between a neuroticism predictive network model with a network predictive of alcohol-use, as well as an impulsivity predictive network model with the alcohol-use risk model. Interestingly, the neuroticism and impulsivity networks were anatomically distinct, sharing only approximately three percent of functional connections (Cheng et al. 2025). These results suggest that both impulsivity and neuroticism appear to share brain commonalities associated with alcohol-use risk and point to potential independent mechanisms that might place youth at risk for issues with drinking. Distinctions have also been found in the brain networks predictive of cocaine and cannabis abstinence (sharing less than one percent of edges; Lichenstein et al. 2023) as well as opioid abstinence and cocaine abstinence (sharing less than one percent of edges; Lichenstein et al. 2021). That these studies failed to find overlap suggests that simply because models are theoretically related does not mean they will share a statistically significant number of edges. This does not invalidate any of the original models (Scheinost et al. 2019). Rather, it reinforces the importance of phenotypic and diagnostic specificity. Despite a theoretical relationship between cognitive processes or clinical conditions, such as impaired attention and autism, different brain regions might be involved. Or a similar neurobiology might indeed be shared, but the dimension under study (in this case, fMRI), might be too coarse to uncover similarities.

We make no claim about fMRI being the best tool to assess the complex architecture of conditions such as autism and ADHD. However, despite shortcomings, it has numerous strengths. To highlight a few: MRI is widely used in both the human and animal neuroscience communities; datasets are increasingly being shared across the world (Poldrack et al. 2013; Poldrack and Gorgolewski 2017), boosting sample sizes and increasing statistical power (Smith and Nichols 2018), especially useful given the intricacies of the processes we seek to understand in neuroscience; and it is non-invasive, again useful when considering its use in participants with neurodevelopmental conditions like autism or ADHD. Future work could apply multimodal techniques to better understand how similarities or differences between predictive models obtained with fMRI relate to markers obtained at different spatiotemporal scales (see ‘Limitations and future directions’ below for more; Lake et al. 2020; Mandino et al. 2025; Mash et al. 2020). In sum, we suggest it is still a valid effort to search for consistencies in predictive network models, and the lack of overlap here across all five models reinforces that phenotypic differences matter. In other words, a parsimonious explanation of the current findings is that network models tend to be specific to the construct—sustained attention or autism traits—they were built to predict.

### Neurobiological patterns observed in transdiagnostic network models

Although perfect overlap was not observed in all five models, transdiagnostic neurobiological patterns emerged. In the positive network, visual, motor, and default mode networks had the highest fraction of edges in the five samples. In the negative network, visual, default mode, and cerebellar networks had the highest fraction of edges. When we considered overlap among the datasets predicting autistic phenotypes, we observed three shared network pairs. All three comprised connections involving medial frontal, default mode, or frontoparietal networks. When we assessed commonalities among the datasets predicting attention phenotypes, we observed three shared network pairs, involving visual, motor, and default mode networks. One interpretation of these findings is that, given the cognitive demands associated with sustaining attention (Fortenbaugh et al. 2017; Esterman and Rothlein 2019), coordination is required between sensory networks and networks mediating higher-order functions, such as the frontoparietal, medial frontal, and default mode network. In addition, the fact that connections linking visual and motor networks emerged as consistently important in the attention samples may reflect the in-scanner button presses associated with the gradCPT.

In addition, our finding that the cerebellum is important in the negative network is intriguing, especially in the context of predicting sustained attention phenotypes (Figure 6D; also see Figure 5 and Supplemental Table 4, with the cerebellum containing the highest degree node across all models). In CPM, edges are assigned to the negative network based on a negative correlation with the phenotypic variable (Shen et al. 2017). Previous work (Marek et al. 2018) suggests that the cerebellum may play a key role in mediating the complex interplay between association networks responsible for higher-order functions, like attention. It is possible that individual differences in within-cerebellum connectivity could be associated with differences in the ability to sustain attention (and in the case of the negative network, greater connectivity is associated with lower ability to sustain attention). Due to the well-known role of the cerebellum in coordinating movement (Buckner et al. 2011), however, and the button pressing aspect of the gradCPT requiring motor coordination, more work is needed to better understand the role of the cerebellum in sustained attention.

In CPM, edges are assigned to the positive network when greater connectivity strength is associated with a positive correlation with a phenotype (Shen et al. 2017). All of the models predicting sustained attention performance (Figure 6D, top matrix) showed the motor-visual II network as being statistically significant. That is, greater connectivity between the networks was associated with better sustained attention. The motor component of the gradCPT is one explanation for this, though it is possible connectivity between these networks helps mediate cognitive functions, such as the ability to sustain attention, as well; impairments in connectivity involving visual to motor networks have been observed in patients with mild cognitive impairments (Yu et al. 2025). For the models predicting autistic traits (Figure 6C, top matrix), connections linking the frontoparietal and medial frontal networks were statistically significant in the positive network. Given the importance of connectivity within and between these networks in mediating demands such as interacting with others in a social context (Yerys et al. 2015; Schurz et al. 2020; Feng et al. 2021), our results are generally aligned with the literature, especially considering that social difficulties are a hallmark of an autism diagnosis (American Psychiatric Association, 2013).

### Factors associated with brain-behavior relationships in the current work

In the RSA, run type, behavioral measure, age, and autism diagnosis were not statistically significant predictors of model similarity. Nevertheless, we did find that when phenotypes were congruent, edge-behavior vectors tended to be more similar (Figure 7) and vice-versa. One interpretation is that while a variable like behavioral measure does not perfectly modulate brain-sustained attention relationships across all five of the samples, there are still instances in which two vectors will be similar because they predicted similar phenotypes (i.e., there is still phenotypic specificity, but the phenotypes affect the models differently, precluding a relationship among all five models considered here).

Another possibility for the seemingly discrepant findings regarding the effect of behavioral measure is that there are other confounding factors that were unmodelled. We restricted analyses to demographic features that were common to all five datasets. Future work in prospective studies could more rigorously assess other variables. Given the importance of other sociodemographic factors in brain-behavior models (Greene et al. 2022; Ricard et al. 2023), particular focus could be placed on influences such as socioeconomic status (Yang et al. 2025), air pollution (Kusters et al. 2025), and other environmental exposures (e.g., Lichenstein et al. 2025).

### Limitations and future directions

There are a number of limitations and future directions to note. We assessed predictive models generated using CPM. Future work could study other modelling methods. In addition, the published models examined here all relied on edge selection to generate binary network masks. We accounted for this by ensuring all networks were of the same size, as well as returning to the original unthresholded data of each study. Nevertheless, there are alternative ways to summarize model weights (Ryali et al. 2012; Smith et al. 2015; He et al. 2020; Ooi et al. 2022; Shafiei et al. 2024) other than the feature selection method described here. In fact, recent work suggests that edges often ignored by feature selection still retain predictive utility (Adkinson et al. 2026). Other groups could investigate the extent to which other feature selection methods are consistent and offer interpretable neurobiological insights.

The samples used in the original studies are somewhat small, particularly when compared with samples such as the Adolescent Brain Cognitive Development (ABCD; Casey et al. 2018) Study or the UK Biobank (Miller et al. 2016). In larger samples, the improved statistical power would improve the ability to more robustly determine feature importance (Marek et al. 2022), especially in heterogeneous conditions like autism (Tang et al. 2020; Pagani et al. 2026).

Longitudinal study designs could be used as well to help determine the impact of intra-subject reliability on model overlap. Assessing whether commonalities can be found from other imaging modalities, such as structural predictive models (Yang et al. 2013), remains an area of further study. There has been growing interest in using precision functional mapping methods (Gordon et al. 2017). Future work could examine consistencies in predictive models generated using longer scan times. Other future research involving animal models and invasive techniques could use multi-modal approaches to better appreciate how similarities or differences at one level of analysis (fMRI) relate to underlying cellular differences (e.g., obtained through wide-field Ca^2+^ imaging) (Lake et al. 2020; Mandino et al. 2025). Similar multi-modal approaches could be used in humans to better appreciate how predictive network fMRI models of autism or attention relate to markers obtained using EEG simultaneously (e.g., Mash et al. 2020).

## Conclusion

We have characterized neurobiological trends in predictive models of sustained attention and autism symptoms in transdiagnostic samples comprising autism and neurotypical participants. Models predicting attention phenotypes shared more similar features with each other than with models predicting autism symptoms and vice-versa. Functional connectivity patterns associated with attention and autism symptoms showed little similarity when considered across phenotypes, and factors such as run type, participant age, and other demographic factors did not impact model similarities. In sum, our data suggest that functional connectivity patterns predicting individual differences in behavior tend to be phenotype-specific, regardless of age or clinical diagnosis.

## Data Availability

Preprocessing of the original datasets (as well as glass brain and circle plot visualizations) used freely available software: https://medicine.yale.edu/bioimaging/suite/. CPM code used to generate the original predictive models is available here: https://github.com/YaleMRRC/CPM. Template scripts used for preprocessing and analysis of the functional data are available here, as are the node-to-network labels: https://github.com/clhorien/transdiagnostic_attn_overlap/tree/main. The functional parcellation is available here: https://www.nitrc.org/frs/?group_id=51. ABIDE data are available here: https://fcon_1000.projects.nitrc.org/indi/abide/. Data from the avCPT sample are available here: https://osf.io/bt2xy/overview. All other data or materials are available from the authors upon request.

## Acknowledgements

This work was supported by a Penn Psychiatry Residency Research Track award R25MH119043-07 (CH) and NIH grant P50MH115716. A.S.K. was supported by a NARSAD Young Investigator Award from the Brain and Behavior Research Foundation (BBRF). E.B.B. was supported by NIMH grant (K23MH133118) and the BBRF (31319).

## References

Adkinson BD, Rosenblatt M, Sun H, Dadashkarimi J, Tejavibulya L, Horien C, Westwater ML, Rodriguez RX, Noble S, Scheinost D. 2026. Feature selection leads to divergent neurobiological interpretations of brain-based machine learning biomarkers. Nat Hum Behav.

Allen G, Courchesne E. 2001. Attention function and dysfunction in autism. Front Biosci. 6:D105–119.

American Psychiatric Association. 2013. Diagnostic and statistical manual of mental disorders (5th ed.). American Psychiatric Association.

Ayano G, Demelash S, Gizachew Y, Tsegay L, Alati R. 2023. The global prevalence of attention deficit hyperactivity disorder in children and adolescents: An umbrella review of meta-analyses. J Affect Disord. 339:860–866.

Beaty RE, Kenett YN, Christensen AP, Rosenberg MD, Benedek M, Chen Q, Fink A, Qiu J, Kwapil TR, Kane MJ, Silvia PJ. 2018. Robust prediction of individual creative ability from brain functional connectivity. Proc Natl Acad Sci U S A. 115:1087–1092.

Benjamini Y, Hochberg, Y. 1995. Controlling the false discovery rate: a practical and powerful approach to multiple testing. Journal of the Royal Statistical Society, Series B. 57:289–300.

Bland JM, Altman DG. 1995. Multiple significance tests: the Bonferroni method. BMJ. 310:170.

Bonnelle V, Leech R, Kinnunen KM, Ham TE, Beckmann CF, De Boissezon X, Greenwood RJ, Sharp DJ. 2011. Default mode network connectivity predicts sustained attention deficits after traumatic brain injury. J Neurosci. 31:13442–13451.

Brereton AV, Tonge BJ, Einfeld SL. 2006. Psychopathology in children and adolescents with autism compared to young people with intellectual disability. J Autism Dev Disord. 36:863–870.

Buckner RL, Krienen FM, Castellanos A, Diaz JC, Yeo BT. 2011. The organization of the human cerebellum estimated by intrinsic functional connectivity. J Neurophysiol. 106:2322–2345.

Cai Y, Mai Z, Li M, Zhou X, Ma N. 2021. Altered frontal connectivity after sleep deprivation predicts sustained attentional impairment: A resting-state functional magnetic resonance imaging study. J Sleep Res. 30:e13329.

Casey BJ, Cannonier T, Conley MI, Cohen AO, Barch DM, Heitzeg MM, Soules ME, Teslovich T, Dellarco DV, Garavan H, Orr CA, Wager TD, Banich MT, Speer NK, Sutherland MT, Riedel MC, Dick AS, Bjork JM, Thomas KM, Chaarani B, Mejia MH, Hagler DJ, Jr., Daniela Cornejo M, Sicat CS, Harms MP, Dosenbach NUF, Rosenberg M, Earl E, Bartsch H, Watts R, Polimeni JR, Kuperman JM, Fair DA, Dale AM, Workgroup AIA. 2018. The Adolescent Brain Cognitive Development (ABCD) study: Imaging acquisition across 21 sites. Dev Cogn Neurosci. 32:43–54.

Cheng A, Lichenstein S, Chaarani B, Liang Q, Babaeianjelodar M, Riley SJ, Luo W, Horien C, Greene AS, Banaschewski T, Bokde ALW, Desrivieres S, Flor H, Grigis A, Gowland P, Heinz A, Bruhl R, Martinot JL, Martinot MP, Artiges E, Nees F, Papadopoulos Orfanos D, Poustka L, Hohmann S, Holz N, Baeuchl C, Smolka MN, Vaidya N, Walter H, Whelan R, Schumann G, Constable RT, Pearlson G, Garavan H, Yip SW. 2025. Impulsivity and neuroticism share distinct functional connectivity signatures with alcohol-use risk in youth. Mol Psychiatry.

Christakou A, Murphy CM, Chantiluke K, Cubillo AI, Smith AB, Giampietro V, Daly E, Ecker C, Robertson D, consortium MA, Murphy DG, Rubia K. 2013. Disorder-specific functional abnormalities during sustained attention in youth with Attention Deficit Hyperactivity Disorder (ADHD) and with autism. Mol Psychiatry. 18:236–244.

Constantino JN, Davis SA, Todd RD, Schindler MK, Gross MM, Brophy SL, Metzger LM, Shoushtari CS, Splinter R, Reich W. 2003. Validation of a brief quantitative measure of autistic traits: comparison of the social responsiveness scale with the autism diagnostic interview-revised. J Autism Dev Disord. 33:427–433.

Corbett BA, Constantine LJ. 2006. Autism and attention deficit hyperactivity disorder: assessing attention and response control with the integrated visual and auditory continuous performance test. Child Neuropsychol. 12:335–348.

Corbett BA, Constantine LJ, Hendren R, Rocke D, Ozonoff S. 2009. Examining executive functioning in children with autism spectrum disorder, attention deficit hyperactivity disorder and typical development. Psychiatry Res. 166:210–222.

Corriveau A, Ke J, Terashima H, Kondo HM, Rosenberg MD. 2025. Functional brain networks predicting sustained attention are not specific to perceptual modality. Netw Neurosci. 9:303–325.

Cortese S, Aoki YY, Itahashi T, Castellanos FX, Eickhoff SB. 2021. Systematic Review and Meta-analysis: Resting-State Functional Magnetic Resonance Imaging Studies of Attention-Deficit/Hyperactivity Disorder. J Am Acad Child Adolesc Psychiatry. 60:61–75.

Cox RW. 1996. AFNI: software for analysis and visualization of functional magnetic resonance neuroimages. Comput Biomed Res. 29:162–173.

Danielson ML, Claussen AH, Bitsko RH, Katz SM, Newsome K, Blumberg SJ, Kogan MD, Ghandour R. 2024. ADHD Prevalence Among U.S. Children and Adolescents in 2022: Diagnosis, Severity, Co-Occurring Disorders, and Treatment. J Clin Child Adolesc Psychol. 53:343–360.

Di Martino A, O’Connor D, Chen B, Alaerts K, Anderson JS, Assaf M, Balsters JH, Baxter L, Beggiato A, Bernaerts S, Blanken LME, Bookheimer SY, Braden BB, Byrge L, Castellanos FX, Dapretto M, Delorme R, Fair DA, Fishman I, Fitzgerald J, Gallagher L, Keehn RJJ, Kennedy DP, Lainhart JE, Luna B, Mostofsky SH, Muller RA, Nebel MB, Nigg JT, O’Hearn K, Solomon M, Toro R, Vaidya CJ, Wenderoth N, White T, Craddock RC, Lord C, Leventhal B, Milham MP. 2017. Data Descriptor: Enhancing studies of the connectome in autism using the autism brain imaging data exchange II. Sci Data. 4.

Di Martino A, Yan CG, Li Q, Denio E, Castellanos FX, Alaerts K, Anderson JS, Assaf M, Bookheimer SY, Dapretto M, Deen B, Delmonte S, Dinstein I, Ertl-Wagner B, Fair DA, Gallagher L, Kennedy DP, Keown CL, Keysers C, Lainhart JE, Lord C, Luna B, Menon V, Minshew NJ, Monk CS, Mueller S, Muller RA, Nebel MB, Nigg JT, O’Hearn K, Pelphrey KA, Peltier SJ, Rudie JD, Sunaert S, Thioux M, Tyszka JM, Uddin LQ, Verhoeven JS, Wenderoth N, Wiggins JL, Mostofsky SH, Milham MP. 2014. The autism brain imaging data exchange: towards a large-scale evaluation of the intrinsic brain architecture in autism. Mol Psychiatr. 19:659–667.

Di Martino A, Zuo XN, Kelly C, Grzadzinski R, Mennes M, Schvarcz A, Rodman J, Lord C, Castellanos FX, Milham MP. 2013. Shared and distinct intrinsic functional network centrality in autism and attention-deficit/hyperactivity disorder. Biol Psychiatry. 74:623–632.

DuPaul GJ. 1998. ADHD rating scale-IV : checklists, norms, and clinical interpretation. New York: Guilford Press.

Esterman M, Noonan SK, Rosenberg M, DeGutis J. 2013. In the Zone or Zoning Out? Tracking Behavioral and Neural Fluctuations During Sustained Attention. Cereb Cortex. 23:2712–2723.

Esterman M, Rothlein D. 2019. Models of sustained attention. Curr Opin Psychol. 29:174–180.

Facoetti A, Paganoni P, Turatto M, Marzola V, Mascetti GG. 2000. Visual-spatial attention in developmental dyslexia. Cortex. 36:109–123.

Feng C, Eickhoff SB, Li T, Wang L, Becker B, Camilleri JA, Hetu S, Luo Y. 2021. Common brain networks underlying human social interactions: Evidence from large-scale neuroimaging meta-analysis. Neurosci Biobehav Rev. 126:289–303.

Feng Q, Ren Z, Wei D, Liu C, Wang X, Li X, Tie B, Tang S, Qiu J. 2024. Connectome-based predictive modeling of Internet addiction symptomatology. Soc Cogn Affect Neurosci. 19.

Finn ES, Shen X, Scheinost D, Rosenberg MD, Huang J, Chun MM, Papademetris X, Constable RT. 2015. Functional connectome fingerprinting: identifying individuals using patterns of brain connectivity. Nat Neurosci. 18:1664–1671.

Fortenbaugh FC, DeGutis J, Esterman M. 2017. Recent theoretical, neural, and clinical advances in sustained attention research. Ann N Y Acad Sci. 1396:70–91.

Garretson HB, Fein D, Waterhouse L. 1990. Sustained attention in children with autism. J Autism Dev Disord. 20:101–114.

Global Burden of Disease Study Autism Spectrum. 2025 (Santomauro et al.). The global epidemiology and health burden of the autism spectrum: findings from the Global Burden of Disease Study 2021. Lancet Psychiatry. 12:111–121.

Gordon EM, Laumann TO, Gilmore AW, Newbold DJ, Greene DJ, Berg JJ, Ortega M, Hoyt-Drazen C, Gratton C, Sun H, Hampton JM, Coalson RS, Nguyen AL, McDermott KB, Shimony JS, Snyder AZ, Schlaggar BL, Petersen SE, Nelson SM, Dosenbach NUF. 2017. Precision Functional Mapping of Individual Human Brains. Neuron. 95:791–807 e797.

Greene AS, Gao SY, Scheinost D, Constable RT. 2018. Task-induced brain state manipulation improves prediction of individual traits. Nat Commun. 9.

Greene AS, Shen X, Noble S, Horien C, Hahn CA, Arora J, Tokoglu F, Spann MN, Carrion CI, Barron DS, Sanacora G, Srihari VH, Woods SW, Scheinost D, Constable RT. 2022. Brain-phenotype models fail for individuals who defy sample stereotypes. Nature. 609:109–118.

He T, Kong R, Holmes AJ, Nguyen M, Sabuncu MR, Eickhoff SB, Bzdok D, Feng J, Yeo BTT. 2020. Deep neural networks and kernel regression achieve comparable accuracies for functional connectivity prediction of behavior and demographics. Neuroimage. 206:116276.

Horien C, Fontenelle S, Joseph K, Powell N, Nutor C, Fortes D, Butler M, Powell K, Macris D, Lee K, Greene AS, McPartland JC, Volkmar FR, Scheinost D, Chawarska K, Constable RT. 2020. Low-motion fMRI data can be obtained in pediatric participants undergoing a 60-minute scan protocol. Sci Rep-Uk. 10.

Horien C, Greene AS, Shen X, Fortes D, Brennan-Wydra E, Banarjee C, Foster R, Donthireddy V, Butler M, Powell K, Vernetti A, Mandino F, O’Connor D, Lake EMR, McPartland JC, Volkmar FR, Chun M, Chawarska K, Rosenberg MD, Scheinost D, Constable RT. 2023. A generalizable connectome-based marker of in-scan sustained attention in neurodiverse youth. Cereb Cortex. 33:6320–6334.

Horien C, Mandino F, Greene AS, Shen X, Powell K, Vernetti A, O’Connor D, Adkinson BD, Tejavibulya L, McPartland JC, Volkmar FR, Chun M, Chawarska K, Lake EMR, Rosenberg MD, Satterthwaite T, Scheinost D, Finn ES, Constable RT. 2026. Optimizing functional connectivity scanning conditions for predicting autistic traits. Nat Ment Health. 4:792–805.

Horien C, Noble S, Finn ES, Shen X, Scheinost D, Constable RT. 2018. Considering factors affecting the connectome-based identification process: Comment on Waller et al. Neuroimage. 169:172–175.

Horien C, Shen X, Scheinost D, Constable RT. 2019. The individual functional connectome is unique and stable over months to years. Neuroimage. 189:676–687.

Huang-Pollock CL, Karalunas SL, Tam H, Moore AN. 2012. Evaluating vigilance deficits in ADHD: a meta-analysis of CPT performance. J Abnorm Psychol. 121:360–371.

Ingersoll B. 2010. Broader autism phenotype and nonverbal sensitivity: evidence for an association in the general population. J Autism Dev Disord. 40:590–598.

Johnson KA, Robertson IH, Kelly SP, Silk TJ, Barry E, Daibhis A, Watchorn A, Keavey M, Fitzgerald M, Gallagher L, Gill M, Bellgrove MA. 2007. Dissociation in performance of children with ADHD and high-functioning autism on a task of sustained attention. Neuropsychologia. 45:2234–2245.

Joshi A, Scheinost D, Okuda H, Belhachemi D, Murphy I, Staib LH, Papademetris X. 2011. Unified framework for development, deployment and robust testing of neuroimaging algorithms. Neuroinformatics. 9:69–84.

Kardan O, Stier AJ, Cardenas-Iniguez C, Schertz KE, Pruin JC, Deng Y, Chamberlain T, Meredith WJ, Zhang X, Bowman JE, Lakhtakia T, Tindel L, Avery EW, Lin Q, Yoo K, Chun MM, Berman MG, Rosenberg MD. 2022. Differences in the functional brain architecture of sustained attention and working memory in youth and adults. PLoS Biol. 20:e3001938.

Kay BP, Wheelock MD, Siegel JS, Raut RV, Chauvin RJ, Metoki A, Rajesh A, Eck A, Pollaro J, Wang A, Suljic V, Adeyemo B, Baden NJ, Scheidter KM, Monk JS, Whiting FI, Ramirez-Perez N, Krimmel SR, Shinohara RT, Tervo-Clemmens B, Hermosillo RJM, Nelson SM, Hendrickson TJ, Madison T, Moore LA, Miranda-Domínguez Ó, Randolph A, Feczko E, Roland JL, Nicol GE, Laumann TO, Marek S, Gordon EM, Raichle ME, Barch DM, Fair DA, Dosenbach NUF. Stimulant medications affect arousal and reward, not attention networks. Cell. 2025 Dec 24;188(26):7529–7546.e20. doi: 10.1016/j.cell.2025.11.039. PMID: 41448140; PMCID: PMC12834599.

Kernbach JM, Satterthwaite TD, Bassett DS, Smallwood J, Margulies D, Krall S, Shaw P, Varoquaux G, Thirion B, Konrad K, Bzdok D. 2018. Shared endo-phenotypes of default mode dysfunction in attention deficit/hyperactivity disorder and autism spectrum disorder. Transl Psychiatry. 8:133.

Kessler D, Angstadt M, Sripada C. 2016. Growth Charting of Brain Connectivity Networks and the Identification of Attention Impairment in Youth. JAMA Psychiat. 73:481–489.

Kriegeskorte N, Mur M, Bandettini P. 2008. Representational similarity analysis - connecting the branches of systems neuroscience. Front Syst Neurosci. 2:4.

Kusters MSW, Granes L, Petricola S, Tiemeier H, Muetzel RL, Guxens M. 2025. Exposure to residential air pollution and the development of functional connectivity of brain networks throughout adolescence. Environ Int. 196:109245.

Lake EMR, Finn ES, Noble SM, Vanderwal T, Shen X, Rosenberg MD, Spann MN, Chun MM, Scheinost D, Constable RT. 2019. The Functional Brain Organization of an Individual Allows Prediction of Measures of Social Abilities Transdiagnostically in Autism and Attention-Deficit/Hyperactivity Disorder. Biol Psychiatry. 86:315–326.

Lake EMR, Ge X, Shen X, Herman P, Hyder F, Cardin JA, Higley MJ, Scheinost D, Papademetris X, Crair MC, Constable RT. 2020. Simultaneous cortex-wide fluorescence Ca(2+) imaging and whole-brain fMRI. Nat Methods. 17:1262–1271.

Landry O, Parker A. 2013. A meta-analysis of visual orienting in autism. Front Hum Neurosci. 7:833.

Lau-Zhu A, Fritz A, McLoughlin G. 2019. Overlaps and distinctions between attention deficit/hyperactivity disorder and autism spectrum disorder in young adulthood: Systematic review and guiding framework for EEG-imaging research. Neurosci Biobehav Rev. 96:93–115.

Lichenstein SD, Kohler R, Ye F, Potenza MN, Kiluk B, Yip SW. 2023. Distinct neural networks predict cocaine versus cannabis treatment outcomes. Mol Psychiatry. 28:3365–3372.

Lichenstein SD, Scheinost D, Potenza MN, Carroll KM, Yip SW. 2021. Dissociable neural substrates of opioid and cocaine use identified via connectome-based modelling. Mol Psychiatry. 26:4383–4393.

Lichenstein SD, Weng Y, Robinson H, Rodriguez L, Babaeianjelodar M, Maynard J, Metayer M, Suneja S, Ojha A, Horien C, Greene AS, Constable RT, Moore TM, Barzilay R, Yip SW, Keller AS. Multivariate environmental exposures are reflected in whole-brain functional connectivity and cognition in youth. Dev Cogn Neurosci. 2026 Oct;81:101788. doi: 10.1016/j.dcn.2026.101788. Epub 2026 Jul 22.

Lin, YJ., Chiu, YN., Wu, YY. et al. Developmental Changes of Autistic Symptoms, ADHD Symptoms, and Attentional Performance in Children and Adolescents with Autism Spectrum Disorder. J Autism Dev Disord 53, 2555–2569 (2023). 10.1007/s10803-022-05445-x

Lord C RM, DiLavore PC, Risi S, Gotham K, Bishop S. 2012. Autism Diagnostic Observation Schedule, Second Edition. Torrance, CA: Western Psychological Services.

Lukito S, Norman L, Carlisi C, Radua J, Hart H, Simonoff E, Rubia K. 2020. Comparative meta-analyses of brain structural and functional abnormalities during cognitive control in attention-deficit/hyperactivity disorder and autism spectrum disorder. Psychol Med. 50:894–919.

Lyall K, Schweitzer JB, Schmidt RJ, Hertz-Picciotto I, Solomon M. 2017. Inattention and hyperactivity in association with autism spectrum disorders in the CHARGE study. Res Autism Spectr Disord. 35:1–12.

Mandino F, Horien C, Shen X, Desrosiers-Gregoire G, Luo W, Markicevic M, Constable RT, Papademetris X, Chakravarty MM, Betzel RF, Lake EMR. 2025. Multimodal identification of the mouse brain using simultaneous Ca(2+) imaging and fMRI. Commun Biol. 8:665.

Marek S, Siegel JS, Gordon EM, Raut RV, Gratton C, Newbold DJ, Ortega M, Laumann TO, Adeyemo B, Miller DB, Zheng A, Lopez KC, Berg JJ, Coalson RS, Nguyen AL, Dierker D, Van AN, Hoyt CR, McDermott KB, Norris SA, Shimony JS, Snyder AZ, Nelson SM, Barch DM, Schlaggar BL, Raichle ME, Petersen SE, Greene DJ, Dosenbach NUF. 2018. Spatial and Temporal Organization of the Individual Human Cerebellum. Neuron. 100:977–993 e977.

Marek S, Tervo-Clemmens B, Calabro FJ, Montez DF, Kay BP, Hatoum AS, Donohue MR, Foran W, Miller RL, Hendrickson TJ, Malone SM, Kandala S, Feczko E, Miranda-Dominguez O, Graham AM, Earl EA, Perrone AJ, Cordova M, Doyle O, Moore LA, Conan GM, Uriarte J, Snider K, Lynch BJ, Wilgenbusch JC, Pengo T, Tam A, Chen J, Newbold DJ, Zheng A, Seider NA, Van AN, Metoki A, Chauvin RJ, Laumann TO, Greene DJ, Petersen SE, Garavan H, Thompson WK, Nichols TE, Yeo BTT, Barch DM, Luna B, Fair DA, Dosenbach NUF. 2022. Reproducible brain-wide association studies require thousands of individuals. Nature. 603:654–660.

Mash LE, Keehn B, Linke AC, Liu TT, Helm JL, Haist F, Townsend J, Muller RA. 2020. Atypical Relationships Between Spontaneous EEG and fMRI Activity in Autism. Brain Connect. 10:18–28.

Miller KL, Alfaro-Almagro F, Bangerter NK, Thomas DL, Yacoub E, Xu J, Bartsch AJ, Jbabdi S, Sotiropoulos SN, Andersson JL, Griffanti L, Douaud G, Okell TW, Weale P, Dragonu I, Garratt S, Hudson S, Collins R, Jenkinson M, Matthews PM, Smith SM. 2016. Multimodal population brain imaging in the UK Biobank prospective epidemiological study. Nat Neurosci. 19:1523–1536.

Noble S, Scheinost D, Constable RT. 2019. A decade of test-retest reliability of functional connectivity: A systematic review and meta-analysis. Neuroimage. 203:116157.

Noble S, Spann MN, Tokoglu F, Shen X, Constable RT, Scheinost D. 2017. Influences on the Test-Retest Reliability of Functional Connectivity MRI and its Relationship with Behavioral Utility. Cereb Cortex. 27:5415–5429.

O’Halloran L, Cao Z, Ruddy K, Jollans L, Albaugh MD, Aleni A, Potter AS, Vahey N, Banaschewski T, Hohmann S, Bokde ALW, Bromberg U, Buchel C, Quinlan EB, Desrivieres S, Flor H, Frouin V, Gowland P, Heinz A, Ittermann B, Nees F, Orfanos DP, Paus T, Smolka MN, Walter H, Schumann G, Garavan H, Kelly C, Whelan R. 2018. Neural circuitry underlying sustained attention in healthy adolescents and in ADHD symptomatology. Neuroimage. 169:395–406.

Ooi LQR, Chen J, Zhang S, Kong R, Tam A, Li J, Dhamala E, Zhou JH, Holmes AJ, Yeo BTT. 2022. Comparison of individualized behavioral predictions across anatomical, diffusion and functional connectivity MRI. Neuroimage. 263:119636.

Pagani M, Zerbi V, Gini S, Alvino FG, Banerjee A, Barberis A, Basson MA, Bozzi Y, Galbusera A, Ellegood J, Fagiolini M, Lerch JP, Matteoli M, Montani C, Pozzi D, Provenzano G, Scattoni ML, Wenderoth N, Xu T, Lombardo MV, Milham MP, Di Martino A, Gozzi A. 2026. Autism subtypes identified using cross-species functional connectivity analyses. Nat Neurosci. 29:1476–1487.

Poldrack RA, Barch DM, Mitchell JP, Wager TD, Wagner AD, Devlin JT, Cumba C, Koyejo O, Milham MP. 2013. Toward open sharing of task-based fMRI data: the OpenfMRI project. Front Neuroinform. 7:12.

Poldrack RA, Gorgolewski KJ. 2017. OpenfMRI: Open sharing of task fMRI data. Neuroimage. 144:259–261.

Ramírez Echeverría M, Schoo C. [Updated 2022 Nov 19]. Delirium. In. StatPearls [Internet] Treasure Island (FL): StatPearls Publishing.

Rapuano KM, Rosenberg MD, Maza MT, Dennis NJ, Dorji M, Greene AS, Horien C, Scheinost D, Todd Constable R, Casey BJ. 2020. Behavioral and brain signatures of substance use vulnerability in childhood. Dev Cogn Neurosci. 46:100878.

Ricard JA, Parker TC, Dhamala E, Kwasa J, Allsop A, Holmes AJ. 2023. Confronting racially exclusionary practices in the acquisition and analyses of neuroimaging data. Nat Neurosci. 26:4–11.

Rohr CS, Vinette SA, Parsons KAL, Cho IYK, Dimond D, Benischek A, Lebel C, Dewey D, Bray S. 2017. Functional Connectivity of the Dorsal Attention Network Predicts Selective Attention in 4-7 year-old Girls. Cereb Cortex. 27:4350–4360.

Rosenberg M, Noonan S, DeGutis J, Esterman M. 2013. Sustaining visual attention in the face of distraction: a novel gradual-onset continuous performance task. Atten Percept Psycho. 75:426–439.

Rosenberg MD, Chun MM. 2020. Network models of attention and working memory. In: Poeppel D, Mangun, G. R., and M. S. Gazzaniga., editor. The Cognitive Neurosciences MIT Press p pp.311–322.

Rosenberg MD, Finn ES, Scheinost D, Papademetris X, Shen X, Constable RT, Chun MM. 2016. A neuromarker of sustained attention from whole-brain functional connectivity. Nat Neurosci. 19:165–171.

Rosenberg MD, Scheinost D, Greene AS, Avery EW, Kwon YH, Finn ES, Ramani R, Qiu M, Constable RT, Chun MM. 2020. Functional connectivity predicts changes in attention observed across minutes, days, and months. Proc Natl Acad Sci U S A. 117:3797–3807.

Rubia K, Halari R, Cubillo A, Mohammad AM, Brammer M, Taylor E. 2009. Methylphenidate normalises activation and functional connectivity deficits in attention and motivation networks in medication-naive children with ADHD during a rewarded continuous performance task. Neuropharmacology. 57:640–652.

Ryali S, Chen T, Supekar K, Menon V. 2012. Estimation of functional connectivity in fMRI data using stability selection-based sparse partial correlation with elastic net penalty. Neuroimage. 59:3852–3861.

Satterthwaite TD, Elliott MA, Gerraty RT, Ruparel K, Loughead J, Calkins ME, Eickhoff SB, Hakonarson H, Gur RC, Gur RE, Wolf DH. 2013. An improved framework for confound regression and filtering for control of motion artifact in the preprocessing of resting-state functional connectivity data. Neuroimage. 64:240–256.

Scheinost D, Noble S, Horien C, Greene AS, Lake EMR, Salehi M, Gao SY, Shen XL, O’Connor D, Barron DS, Yip SW, Rosenberg MD, Constable RT. 2019. Ten simple rules for predictive modeling of individual differences in neuroimaging. Neuroimage. 193:35–45.

Schurz M, Maliske L, Kanske P. 2020. Cross-network interactions in social cognition: A review of findings on task related brain activation and connectivity. Cortex. 130:142–157.

Seeburger DT, Xu N, Ma M, Larson S, Godwin C, Keilholz SD, Schumacher EH. 2024. Time-varying functional connectivity predicts fluctuations in sustained attention in a serial tapping task. Cogn Affect Behav Neurosci. 24:111–125.

Shafiei G, Keller AS, Bertolero M, Shanmugan S, Bassett DS, Chen AA, Covitz S, Houghton A, Luo A, Mehta K, Salo T, Shinohara RT, Fair D, Hallquist MN, Satterthwaite TD. 2024.

Generalizable Links Between Borderline Personality Traits and Functional Connectivity. Biol Psychiatry. 96:486–494.

Shaw KA Willams S, Patrick ME, et al. 2025. Prevalence and Early Identification of Autism Spectrum Disorder Among Children Aged 4 and 8 Years — Autism and Developmental Disabilities Monitoring Network, 16 Sites, United States, 2022. MMWR Surveill Summ.

Shaywitz SE, Shaywitz BA. 2008. Paying attention to reading: the neurobiology of reading and dyslexia. Dev Psychopathol. 20:1329–1349.

Shen X, Finn ES, Scheinost D, Rosenberg MD, Chun MM, Papademetris X, Constable RT. 2017. Using connectome-based predictive modeling to predict individual behavior from brain connectivity. Nat Protoc. 12:506–518.

Shen X, Tokoglu F, Papademetris X, Constable RT. 2013. Groupwise whole-brain parcellation from resting-state fMRI data for network node identification. Neuroimage. 82:403–415.

Smith SM, Nichols TE. 2018. Statistical Challenges in “Big Data” Human Neuroimaging. Neuron. 97:263–268.

Smith SM, Nichols TE, Vidaurre D, Winkler AM, Behrens TE, Glasser MF, Ugurbil K, Barch DM, Van Essen DC, Miller KL. 2015. A positive-negative mode of population covariation links brain connectivity, demographics and behavior. Nat Neurosci. 18:1565–1567.

Tamon H, Fujino J, Itahashi T, Frahm L, Parlatini V, Aoki YY, Castellanos FX, Eickhoff SB, Cortese S. 2024. Shared and Specific Neural Correlates of Attention Deficit Hyperactivity Disorder and Autism Spectrum Disorder: A Meta-Analysis of 243 Task-Based Functional MRI Studies. Am J Psychiatry. 181:541–552.

Tang SY, Sun NB, Floris DL, Zhang XM, Di Martino A, Yeo BTT. 2020. Reconciling Dimensional and Categorical Models of Autism Heterogeneity: A Brain Connectomics and Behavioral Study. Biol Psychiat. 87:1071–1082.

Thompson GJ, Magnuson ME, Merritt MD, Schwarb H, Pan WJ, McKinley A, Tripp LD, Schumacher EH, Keilholz SD. 2013. Short-time windows of correlation between large-scale functional brain networks predict vigilance intraindividually and interindividually. Hum Brain Mapp. 34:3280–3298.

Yang JJ, Yoon U, Yun HJ, Im K, Choi YY, Lee KH, Park H, Hough MG, Lee JM. 2013. Prediction for human intelligence using morphometric characteristics of cortical surface: partial least square analysis. Neuroscience. 246:351–361.

Yang Y, Kong T, Liu R, Luo L. 2025. Associations of interpersonal and socioeconomic early life adversity dimensions with adolescents’ corticolimbic circuits, cognition, and mental health. Transl Psychiatry. 15:168.

Yerys BE, Gordon EM, Abrams DN, Satterthwaite TD, Weinblatt R, Jankowski KF, Strang J, Kenworthy L, Gaillard WD, Vaidya CJ. 2015. Default mode network segregation and social deficits in autism spectrum disorder: Evidence from non-medicated children. Neuroimage Clin. 9:223–232.

Yu Y, Hu B, Yu XW, Cui YY, Cao XY, Ni MH, Li SN, Dai P, Sun Q, Bai XY, Tong Y, Jing XR, Yang AL, Liang SR, Du LJ, Guo S, Yan LF, Gao B, Cui GB; Alzheimer’s Disease Neuroimaging Initiative. 2025. Dysregulated brain dynamics in the visualmotor network in type 2 diabetes patients and their relationship with cognitive impairment. Brain Res Bull. DOI: 10.1016/j.brainresbull.2025.111313

Zhou G, Xie K, Ngo A, Ding R, Cabalo DG, Wang Y, Smith M, Nakamura M, Royer J, Serediuc N, Toussaint PJ, Mottron L, Di Martino A, Milham MP, Hong S-J, Evans AC, Bernhardt BC. Differential subcortical–cortical circuit reorganization in autism and ADHD. bioRxiv 2026.10.13.758196; doi: 10.64898/2026.10.13.758196

