## Supplemental Materials for "Feature overlap in models of sustained attention and autism symptoms"

**Supplemental Table of Contents**

Supplemental Methods……………………………………………………………………...……………………..3

Full description of datasets used to generate predictive models…………………………………………..3

Preprocessing of imaging data………………………………………………..…………………………...5

Minimizing the impact of head motion ………………………………………………..……………….....6

Supplemental Table 1: Quantifying the number of participants in the Yale autism sample and the youth attention sample with mean FD values below motion thresholds……………..…...…………….7

Functional imaging conditions: avCPT sample (M4).....…………………………………………...……..7

Statistical overlap of edges in predictive models: CDF ………..…………………………………..……..7

Supplemental Figure 1. Previously published models and independent samples used to validate each model……………………………………………………………………………………………………....9

Supplemental Table 2. The CPM and model summarization parameters from each sample in which a CPM network model was generated…....……………………………………………………………......10

Supplemental Figure 2. Connectome-based predictive modelling (CPM) overview figure……………..11

Supplemental Figure 3. Schematic of procedures used to generate nulls………...……………………...12

Supplemental Results………………………………………………………………………………………….....14

Generating summary network models: approach from each publication using CPM……………………………………………………………………………………………………...14

Supplemental Figure 4. Characteristics of previously published predictive models………...…………..15

Supplemental Table 3. Characterizing the connections in the positive and negative networks of previously published models. …………………………………………………………...………........….16

Supplemental Figure 5. Visualizing the previously published predictive models—positive and negative networks……………………...……………………………………………………………………….….17

Supplemental Figure 6. Visualizing the previously published predictive models—additional representations……………………...……………………………………………………………………18

Supplemental Figure 7. Edge occurrences across models using different thresholds*…………………*.….19

Supplemental Figure 8. Edge overlap using different thresholds………………………………………..20

Supplemental Table 4. MNI coordinates and neurobiological context of high degree shared nodes…....21

Supplemental Figure 9. Visualization of edges when models predicted congruent phenotypes: autism models ……………………………………………………………………………………………..…….24

Supplemental Figure 10. Visualization of edges when models predicted congruent phenotypes: attention models ………………………………………………………………………………………………..….25

Supplemental Figure 11. Network matrices using different thresholds……………………...…….…….26

Supplemental Figure 12. Neuroanatomy of the network models: individual datasets and pairwise relationships……………………………………………………………..……….……………..………..28

Supplemental Table 5. Similarity matrix showing similarity of edge-behavior vectors across datasets...29

Supplemental References…………………………………………………………………………………...……30

**Supplemental Methods**

*Full description of datasets used to generate predictive models*

Dataset 1, model 1: ABIDE model (M1)

The Autism Brain Imaging Data Exchange (ABIDE) dataset comprised individuals with and without autism (n = 229, 65 females; mean age = 10.45 years, st. dev. = 1.8 years; mean IQ = 113.7, st. dev. = 15.1; 77/229 participants were diagnosed with autism; Table 1). Throughout the text, we refer to this sample as the ‘ABIDE model/sample’ (Di Martino et al. 2014; Di Martino et al. 2017). Processing of these data is described elsewhere (Lake et al. 2019). Resting-state data were used. Social responsiveness scale (SRS) total scores (reflective of social difficulties in autism; Constantino et al. 2003) were the behavior of interest (mean score = 42.4, st. dev. = 40.2).

Dataset 2, model 2: Yale youth autism model (M2)

The second dataset comprised 63 subjects (mean age = 11.7 years, st. dev. = 2.8 years; 29 females; mean IQ = 107.8, st. dev. = 15.1); full exclusion criteria and imaging parameters have been published elsewhere (Horien et al. 2026). Twenty of the participants had autism; 11 other participants had a different neurodevelopmental condition (five with ADHD, two with anxiety disorder, and four were classified as belonging to the broader autism phenotype) (Ingersoll 2010). Hereafter, we refer to this model/sample as the ‘Yale youth autism model/sample.’ Subjects performed a sustained attention task, the gradual-onset continuous performance task (gradCPT), in the scanner (Esterman et al. 2013; Rosenberg et al. 2013; Rosenberg et al. 2016). Autism symptoms were scored using the Autism Diagnostic Observation Schedule-2 (ADOS-2) (Lord et al. 2012); calibrated total severity scores were used in the present work (mean score = 3.1, st. dev. = 3.1).

Dataset 2, model 3: youth attention model (M3)

The second dataset was also used to generate a different model. The sample comprised 70 subjects (mean age = 11.59 years, st. dev. = 2.87 years; 31 females; mean IQ = 107.23, st. dev. = 15.9); full details of the sample have been described elsewhere (Horien et al. 2020; Horien et al. 2023). Twenty of the participants had autism. Thirteen other participants had a different neurodevelopmental condition (seven with ADHD, two with anxiety disorder, one with bipolar disorder, and three were classified as belonging to the broader autism phenotype) (Ingersoll 2010). We refer to this model/sample as the ‘youth attention model/sample,’ but readers should keep in mind the transdiagnostic nature of the dataset. Subjects performed gradCPT in the scanner (Esterman et al. 2013; Rosenberg et al. 2013; Rosenberg et al. 2016). Accuracy on the gradCPT is the behavior of interest in this sample (*d*’; mean score = 2.54, st. dev. = 0.9). Note that because they were drawn from the same parent studies, models 2 and 3 shared 56 subjects.

Dataset 3, model 4: avCPT model (M4)

The next dataset has been described elsewhere (Corriveau et al. 2025). The final sample size was n = 43 (n = 24 females in visual condition, n = 23 females in auditory condition); average age = 22.4 (st. dev. = 4.18). Participants completed an in-scanner 10-minute audio-visual continuous performance task (avCPT; we refer to this model/sample hereafter as the ‘avCPT model/sample’). Accuracy on the avCPT is the behavior of interest in this sample (*d*’ visual condition: mean score = 3.08, st. dev. = 0.65; *d*’ auditory condition: mean score = 0.95, st. dev. = 0.60).

Dataset 4, model 5: adult attention model (M5)

The last dataset comprised neurotypical adults (hereafter referred to as the ‘adult attention model/sample’; n = 25, 13 females, mean age = 22.8 years, st. dev. = 3.5 years); full details of this sample are described elsewhere (Rosenberg et al. 2016). Subjects performed gradCPT in the scanner (Esterman et al. 2013; Rosenberg et al. 2013; Rosenberg et al. 2016). Accuracy on the gradCPT is the behavior of interest in this sample (*d*’; mean score = 2.11, st. dev. = 0.92).

*Preprocessing of imaging data*

All five original studies used the same general processing pipeline described elsewhere( Greene et al. 2018; Horien et al. 2018; Rapuano et al. 2020; Greene et al. 2022; Corriveau *et al.* 2025). For the Yale youth autism, youth attention, adult attention, and ABIDE samples, preprocessing steps were performed using BioImage Suite (Joshi et al. 2011) unless otherwise noted, and included: skull-stripping and performing linear and non-linear transformations to warp a 268-node functional atlas(Finn et al. 2015) from Montreal Neurological Institute space to single-subject space. Functional images were motion-corrected using SPM8 (<https://www.fil.ion.ucl.ac.uk/spm/software/spm8/>). Covariates of no interest were regressed from the data, including a 24-parameter model of motion (Satterthwaite et al. 2013), mean cerebrospinal fluid signal, mean white matter signal, and the global signal. Data were temporally smoothed with a zero-mean unit-variance low-pass Gaussian filter (approximate cutoff frequency of 0.12 Hz). The only difference in the processing pipelines was the brain extraction tool; optiBET was used in the Yale youth autism and youth attention samples, whereas the BioImage Suite brain extraction tool was used in the ABIDE and adult attention samples.

For the avCPT dataset, data were processed using AFNI (Cox 1996). Briefly, the following steps were performed: removal of leading TRs; alignment of functional data to MNI space; regression of nuisance covariates, (a 24-parameter head motion model: six motion parameters, six temporal derivatives, and their squares; mean signal from subject-level white matter and ventricle masks; and mean global signal); and censoring of frames based on motion (i.e., if the derivative of motion parameters > 0.25 mm or for which > 10% of the brain was outliers).

Note that the original avCPT sample comprised 45 visual and 44 auditory runs; we excluded two participants from the visual run and one participant from the auditory runs for missing a significant number of edges (>4,100 / 35,778 total) due to poor coverage. In the remaining subjects, twelve participants in the auditory runs and 18 subjects in the visual runs had incomplete functional coverage. If any subject was missing an edge, we removed that edge from all participants being compared from all samples; the final connectome size was 32,640 edges and 31,626 edges in the auditory and visual runs, respectively.

*Minimizing the impact of head motion*

In each study, numerous approaches were used to ensure artifacts related to head motion were not driving model results. For instance, in the Yale youth autism and youth attention samples, all participants completed a mock scan protocol approximately a week before the MRI session. In previous work, we have demonstrated that the protocol results in a reduction of in-scanner head motion (Horien *et al.* 2020). In both of these samples this resulted in 100% of participants having functional data below a 0.24 mm mean framewise displacement (FD) threshold (Supplemental Table 1), in line with those used for determining high- versus low-motion data in youth and/or those with a psychiatric condition (Yip et al. 2019; Ju et al. 2020; Lichenstein et al. 2021). The number of participants with a mean FD of < 0.24 mm is lower than other samples of youth(Casey et al. 2018).

In the avCPT (Corriveau *et al.* 2025) sample, censoring was conducted of frames for which the derivative of motion parameters > 0.25 mm or for which more than 10% of the brain were outliers. Runs were excluded if the average FD after censoring > 0.15 mm, if the maximum head displacement > 4 mm, or if > 50% of frames were censored during processing. All final subjects in the avCPT sample comprised functional data < 0.0822 mm mean FD.

In addition, the adult attention and ABIDE samples include low-motion data. Specifically, all subjects from the adult attention sample had a mean FD below 0.087 mm. All subjects from ABIDE had a mean FD below 0.10 mm. Further, numerous control analyses are described in each original study detailing steps taken to demonstrate that head motion is not driving results. We refer the reader to each respective publication for more details (Rosenberg et al. 2016; Lake et al. 2019; Horien et al. 2023; Horien et al. 2026; Corriveau et al. 2025).

|  | Yale youth autism sample (M2) |  | Youth attention sample (M3) |
| --- | --- | --- | --- |
| Motion threshold (mean FD, mm) | Percentage of participants below threshold |  | Percentage of participants below threshold |
| 0.24 | 100% |  | 100% |
| 0.20 | 95.20% |  | 95.70% |
| 0.15 | 88.90% |  | 84.30% |
| 0.10 | 63.50% |  | 61.40% |

Supplemental Table 1. Quantifying the number of participants in the Yale autism sample (left column) and the youth attention sample (right column) with mean FD values below motion thresholds. Data from the youth attention sample are reproduced with permission from (Horien et al. 2023). FD, framewise displacement.

*Functional imaging conditions: avCPT sample (M4)*

Participants in the avCPT sample completed an audio-visual continuous performance task (avCPT) (Corriveau et al. 2025). The task was completed twice in two separate sessions, so that the visual condition required a button press to frequent (90%) visual stimuli and withhold for infrequent (10%) stimuli (indoor and outdoor scenes, counterbalanced across participants); the same frequency parameters were used in the auditory version of the task (natural and artificial sounds, counterbalanced across participants). Performance was quantified using d’ (sensitivity) as above, the participant’s z-scored hit rate minus z-scored false alarm rate.

Statistical overlap of edges in predictive models: CDF

To assess the overlap of edges among specific models themselves in a pairwise fashion, we considered two predictive models at a time and asked if the overlap among edges was statistically significant. To do so, we used the hypergeometric cumulative density function (CDF) as previously described (Rosenberg et al. 2020; Corriveau et al. 2025). Briefly, in MATLAB, the pseudocode is *P* = 1 – hygecdf(*x*, *M*, *K*, *n*), where *x* equals the number of overlapping edges between the two models, *M* equals the total number of edges in the connectome (31,626 edges after removing nodes that were missing from the avCPT visual run), *K* equals the number of edges in the first predictive model being compared, and *n* equals the number of edges in the second predictive model being compared. This allowed us to determine empirical *P*-values of drawing up to *x* of *K* possible items in *n* drawings without replacement from an *M*-item population. We used the Benjamini-Hochberg procedure (Benjamini and Hochberg 1995) to control for multiple comparisons (20 total, ten comparisons among the positive network and ten among the negative networks).

*Previously published models and independent samples used to validate each model*

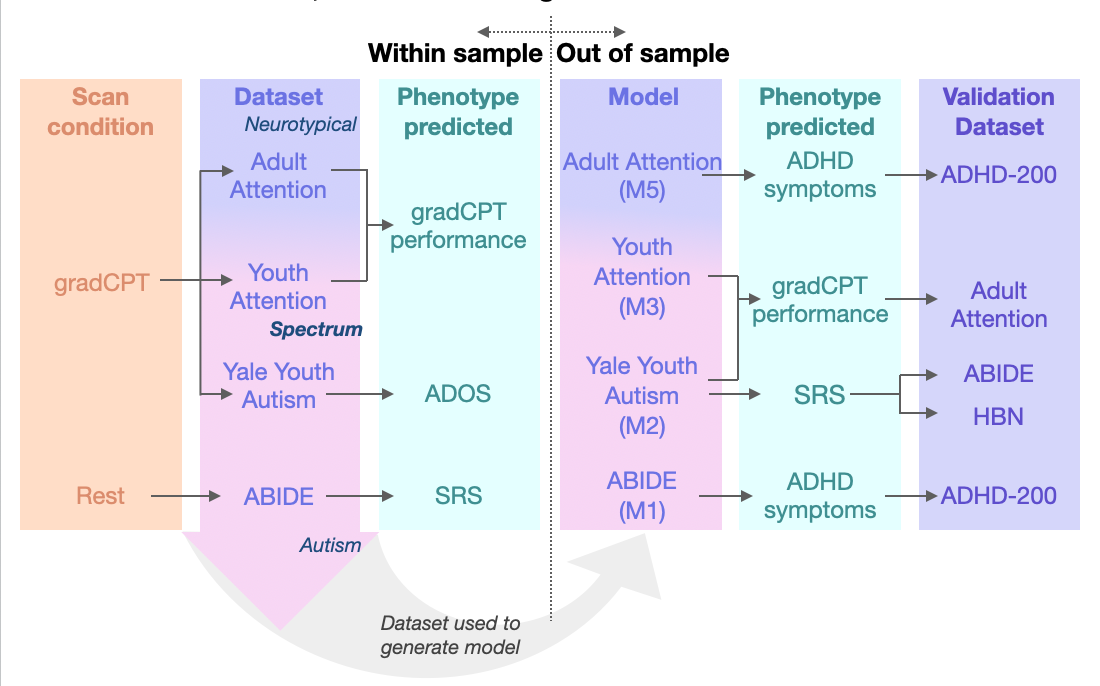

Supplemental Figure 1. Overview of the previously published models. The original sample from which each model was derived, along with the samples used for generalization, are shown. Models were validated via cross-validation in the original dataset listed (‘Within-sample’). The ‘Scan condition’ (gradCPT task or resting state) and the ‘Phenotype predicted’ are shown to the left and right of the ‘Dataset’ column, respectively. The Youth Attention, Yale Youth Autism, and ABIDE datasets all comprised participants with and without autism (indicated by the purple-pink gradient shown in the Dataset column, with pink indicating a higher proportion of participants with autism). The adult attention sample comprised neurotypical participants. After cross-validation, attention models were tested on independent datasets (‘Out of sample’), in some cases on multiple phenotypes, indicated in the ‘Phenotype predicted’ and ‘Validation Dataset’ columns, respectively. Note that the avCPT dataset is not shown here, as the emphasis of the original paper was not on deriving a single model and testing generalizability in external datasets. Rather, it was testing the specificity of multiple models across perceptual modalities. ABIDE, autism brain imaging data exchange; ADHD, attention-deficit/hyperactivity disorder; ADOS, Autism Diagnostic Observation Schedule; gradCPT, gradual-onset continuous performance task; HBN, Healthy Brain Network; M, model; SRS, social responsiveness scale.

|  | Autism models | | Attention models | |
| --- | --- | --- | --- | --- |
|  | ABIDE (M1) | Yale youth autism sample (M2) | Youth attention sample (M3) | Adult attention sample (M5) |
| Cross-validation approach | Leave-one-out | 10-fold | 10-fold | Leave-one-out |
| *P*-value used in edge selection threshold in CPM | 0.01 | 0.05 | 0.05 | 0.01 |
| Model summarization parameters | Edges appearing in at least 3 CPM models trained to predict one of the six SRS subscales | Edges appearing in 6/10 folds and 600/1,000 iterations of CPM | Edges appearing in 6/10 folds and 600/1,000 iterations of CPM | Edges appearing in every fold |

Supplemental Table 2. The CPM and model summarization parameters from each sample in which a CPM network model was generated. Note that the avCPT dataset is not shown here, as the emphasis of the original paper was not on deriving a single model and testing generalizability in external datasets. Rather, it was testing the specificity of multiple models across perceptual modalities. ABIDE, autism brain imaging data exchange; CPM, connectome-based predictive modelling; M, model; SRS, social responsiveness scale.

*Connectome-based predictive modelling (CPM) overview figure*

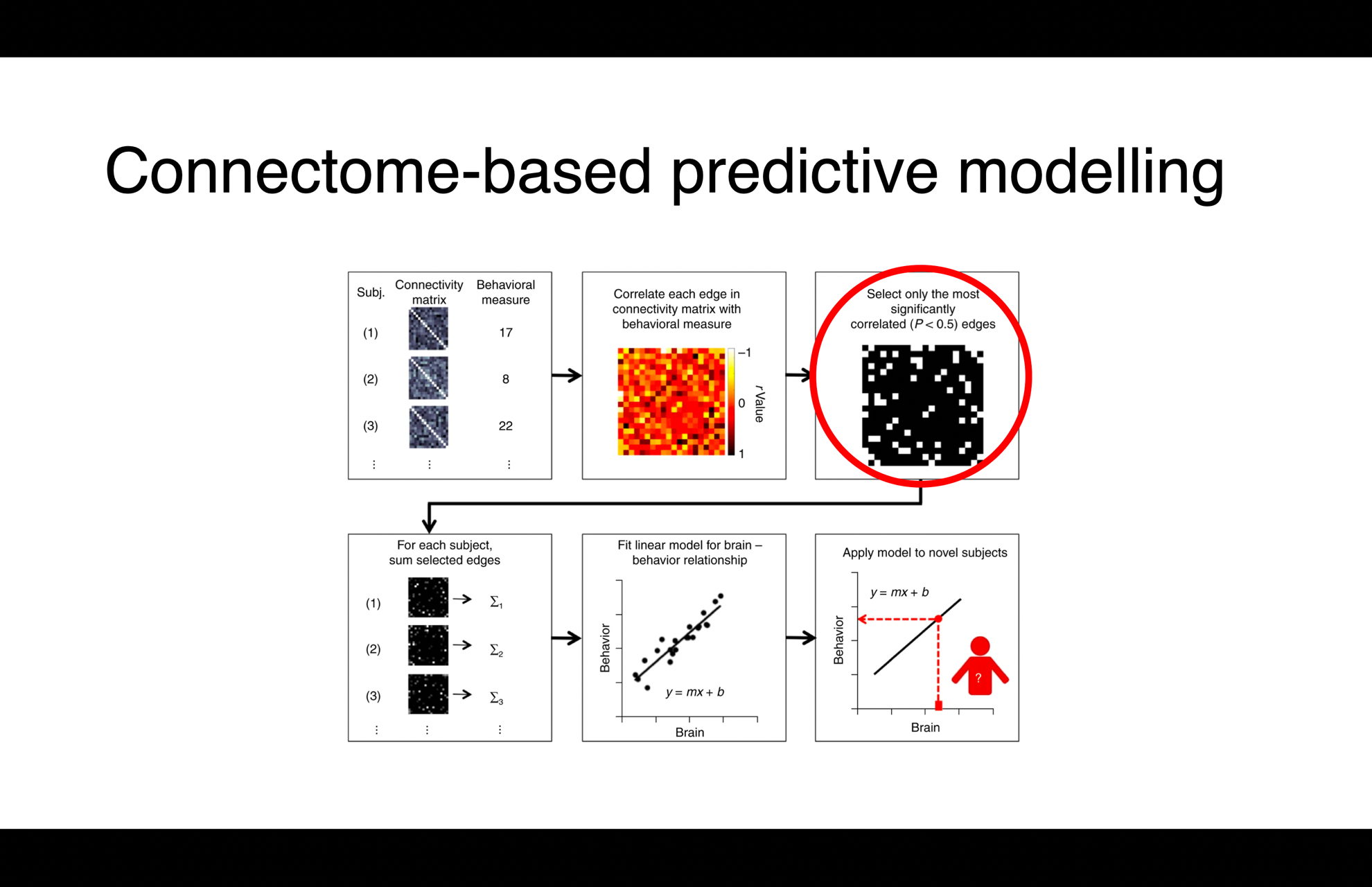

Supplemental Figure 2. A schematic detailing the step-by-step process of CPM. The step leading to network masks is circled in red. However, as specified in ‘Accounting for differences in the size of the predictive models,’ instead of a feature selection step, we retained only the top 1,000 positively and negatively correlated edges for our main analyses. We also tested 500 edges in each positive and negative network (1,000 total), as well as 2,500 edges in each network (5,000 edges total). Figure adapted with permission from Shen et al. (2017).

*Schematic of procedures used to generate nulls*

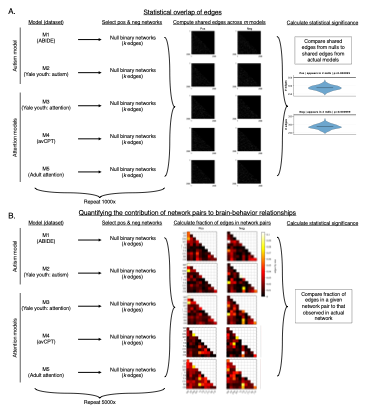

Supplemental Figure 3. Schematic for generating null network models. A) The statistical overlap of edges was assessed via the following procedure. We performed 1,000 iterations of generating random networks, ensuring the null binary networks were the same size as the original network from a given sample (i.e., the number of edges *k* was the same size as that used in the original analysis, 500, 1,000, or 2,500). We then compared the number of times the random networks resulted in a number of overlapping edges greater than or equal to the number of overlapping edges in the actual predictive networks. In other words, we calculated the overlap of random network models and determined how many edges were shared by models. B) The statistical significance of network pairs in the models was assessed via the following procedure. Specifically, we generated random binary networks, with edge locations randomly generated across the connectome, ensuring the size of the null network matched that of the true predictive network from a given dataset (with *k* again equaling 500, 1,000, or 2,500). The process was repeated 5,000 times. We then quantified the number of edges between a given network pair in the null network, correcting for network size. *P*-values were determined by calculating the number of times out of 5,000 null models that a number of edges greater than or equal to the number of edges from the observed network pair from the predictive model were obtained. Given this analysis involved 55 comparisons for a given network, we use the stricter Bonferroni correction (Bland and Altman 1995) when reporting significant *P*-values. ABIDE, Autism Brain Imaging Data exchange; avCPT, audio-visual continuous performance task; M, model; Neg net, negative network; Pos net, positive network. Network labels for the matrices use the same convention as Figure 6 in the main text.

**Supplemental Results**

*Generating summary network models: approach from each publication using CPM*

Slightly different approaches were used to generate summary CPM predictive models from each publication. In general, the approaches involved selecting edges that tended to appear across many cross-validation folds or many iterations of CPM (Supplemental Table 2). In ABIDE (M1), an edge must appear in at least three CPM models trained to predict one of the six SRS subscales. In the Yale youth autism model (M2) and Yale youth attention model (M3), consensus positive and consensus negative networks were defined to include edges that appeared in at least 6/10 folds in 300/500 iterations of CPM. In the adult attention sample (M5), consensus positive and negative networks were defined to include edges that appeared in every fold.

Despite the varied approaches, sizes of the previously published networks were similar. Summing both the number of positive and negative edges resulted in a range of 1,387 – 2,162 total edges across the four models (out of a possible 35,778), representing 3.88 – 6.04% of the total edges in the connectome (Supplemental Figure 4). Approximately 50% of the edges comprised intra-hemispheric connections across the positive and negative networks, with the right and left hemispheres contributing about the same number of edges (Supplemental Table 3). Broadly, all lobes contributed to the models in both the positive and negative networks. Connections were observed within and between cerebellar, subcortical, and cortical areas, both within and across hemispheres (Supplemental Figures 5-6).

*Characteristics of previously published predictive models*

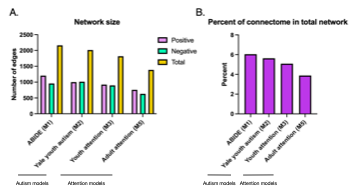

Supplemental Figure 4. The number of edges in different, previously published models of autism traits and sustained attention. A) Barplot showing the number of edges in the positive (purple), negative (green), and the total (i.e., the sum of the positive and negative networks; yellow). The number of edges is shown on the y-axis; the sample from which the model is obtained is shown on the x-axis. B) Barplot showing the percentage of the connectome in each respective model. The total number of edges (corresponding to the yellow bar in A) is divided by the total number of edges in the connectome (35,778). Percent is shown on the y-axis; the sample from which the model is obtained is shown on the x-axis. Note that because we are assessing the models as defined in each publication, we have not accounted for autism by flipping network signs; the positive and negative networks are presented as defined originally. Note also the avCPT dataset is not shown here, as the emphasis of the original paper was not on deriving a model and testing generalizability in external datasets. Rather, it was testing the specificity of multiple models across perceptual modalities. ABIDE, autism brain imaging data exchange.

| Pos net |  | Model | Right Intra | Left Intra | Total Intra | Inter | Total | Percentage Right Intra | Percentage Left Intra | Percentage Total Intra | Percent Inter |
| --- | --- | --- | --- | --- | --- | --- | --- | --- | --- | --- | --- |
|  | Autism models | ABIDE (M1) | 286 | 334 | 620 | 585 | 1205 | 23.7 | 27.7 | 51.5 | 48.5 |
|  |  | Yale youth autism (M2) | 239 | 237 | 476 | 525 | 1001 | 23.9 | 23.7 | 47.6 | 52.4 |
|  | Attention models | Youth Attention (M3) | 252 | 231 | 483 | 439 | 922 | 27.3 | 25.1 | 52.4 | 47.6 |
|  |  | Adult attention (M5) | 199 | 179 | 378 | 379 | 757 | 26.3 | 23.6 | 49.9 | 50.1 |
|  |  | Total | 976 | 981 | 1957 | 1928 | 3885 | 25.1 | 25.3 | 50.4 | 49.6 |
| Neg net | Autism models | ABIDE (M1) | 222 | 231 | 453 | 504 | 957 | 23.2 | 24.1 | 47.3 | 52.7 |
|  |  | Yale youth autism (M2) | 197 | 240 | 437 | 576 | 1013 | 19.4 | 23.7 | 43.1 | 56.9 |
|  | Attention models | Youth Attention (M3) | 252 | 194 | 446 | 450 | 896 | 28.1 | 21.7 | 49.8 | 50.2 |
|  |  | Adult attention (M5) | 144 | 147 | 291 | 339 | 630 | 22.9 | 23.3 | 46.2 | 53.8 |
|  |  | Total | 815 | 812 | 1627 | 1869 | 3496 | 23.3 | 23.2 | 46.5 | 53.5 |

Supplemental Table 3. Characterizing the connections in the positive and negative networks of previously published models. ABIDE, autism brain imaging data exchange; Intra, intra-hemispheric connections; Inter, interhemispheric connections; M, model; Neg net, negative network; Pos net, positive network.

*Visualizing the previously published predictive models—positive and negative networks*

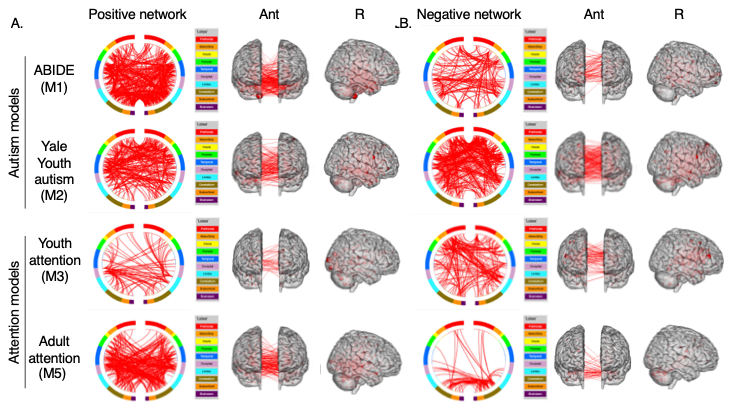

Supplemental Figure 5. The network masks of the CPM networks that have been previously published. A) Positive and B) negative network masks. Each row corresponds to a different network model; the dataset from which the network was obtained is indicated to the left. A circle plot is shown immediately to the right of the dataset name. The topmost aspect of the circle represents anterior; the bottom, posterior. The right half of the circle plot corresponds to the right hemisphere of the brain. A legend indicating the approximate anatomic “lobe” is shown to the right. The same edges are plotted in the glass brains as lines connecting different nodes; in these visualizations, nodes are sized according to degree, the number of edges connected to that node. The view of the brain is indicated at the top of the panel. To aid in visualization in panels A and B, we have thresholded the matrices to only show nodes with a degree threshold > 18. See Supplemental Figure 6A for unthresholded visualizations, as well as the network representations of the models in Supplemental Figure 6B. The connectivity viewer tool of BioImage Suite (<https://medicine.yale.edu/bioimaging/suite/>) was used to generate the circle plots and glass brains shown here. Note that because we are assessing the models as defined in each publication, we have not accounted for autism by flipping network signs; the positive and negative networks are presented as defined originally. Note also the avCPT dataset is not shown here, as the emphasis of the original paper was not on deriving a model and testing generalizability in external datasets. Rather, it was testing the specificity of multiple models across perceptual modalities. ABIDE, autism brain imaging data exchange; Ant, anterior; L, left; M, model; Post, posterior; R, right.

*Visualizing the previously published predictive models—additional representations*

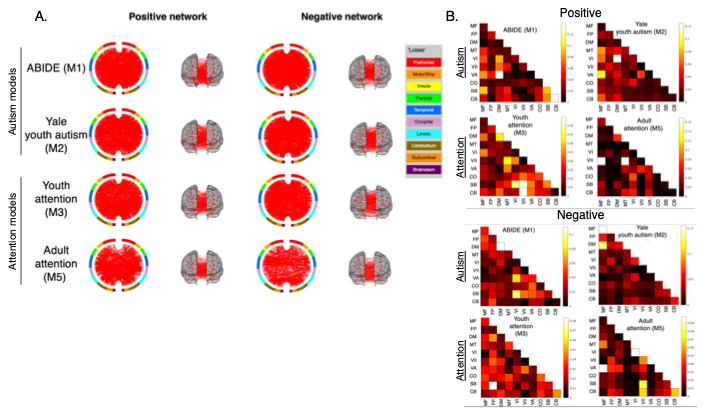

Supplemental Figure 6. Additional representations of the previously published models. A) Unthresholded circle plots and glass brains. Note that these are the same data as shown in Supplemental Figure 5, except no degree threshold has been applied. B) Shown are the network masks for each model. Dataset from which the model was obtained is indicated above each matrix. Each cell in a matrix corresponds to the fraction of edges in a given network pair that were included in the given model, after correcting for network size. The colorbars for each matrix are scaled so that a lower fraction of edges corresponds to darker colors; a higher fraction of edges corresponds to lighter colors. The connectivity viewer tool of BioImage Suite (<https://medicine.yale.edu/bioimaging/suite/>) was used to generate the circle plots and glass brains shown here. Note that because we are assessing the models as defined in each publication, we have not accounted for autism by flipping network signs; the positive and negative networks are presented as defined originally. Note also the avCPT dataset is not shown here, as the emphasis of the original paper was not on deriving a model and testing generalizability in external datasets. Rather, it was testing the specificity of multiple models across perceptual modalities. ABIDE, autism brain imaging data exchange; M, model. Network labels: MF, medial frontal; FP, frontoparietal; DM, default mode; MT, motor; VI, visual I; VII, visual II; VA, visual association; CO, cingulo-opercular; SB, subcortical; CB, cerebellum.

*Edge occurrences across models using different thresholds*

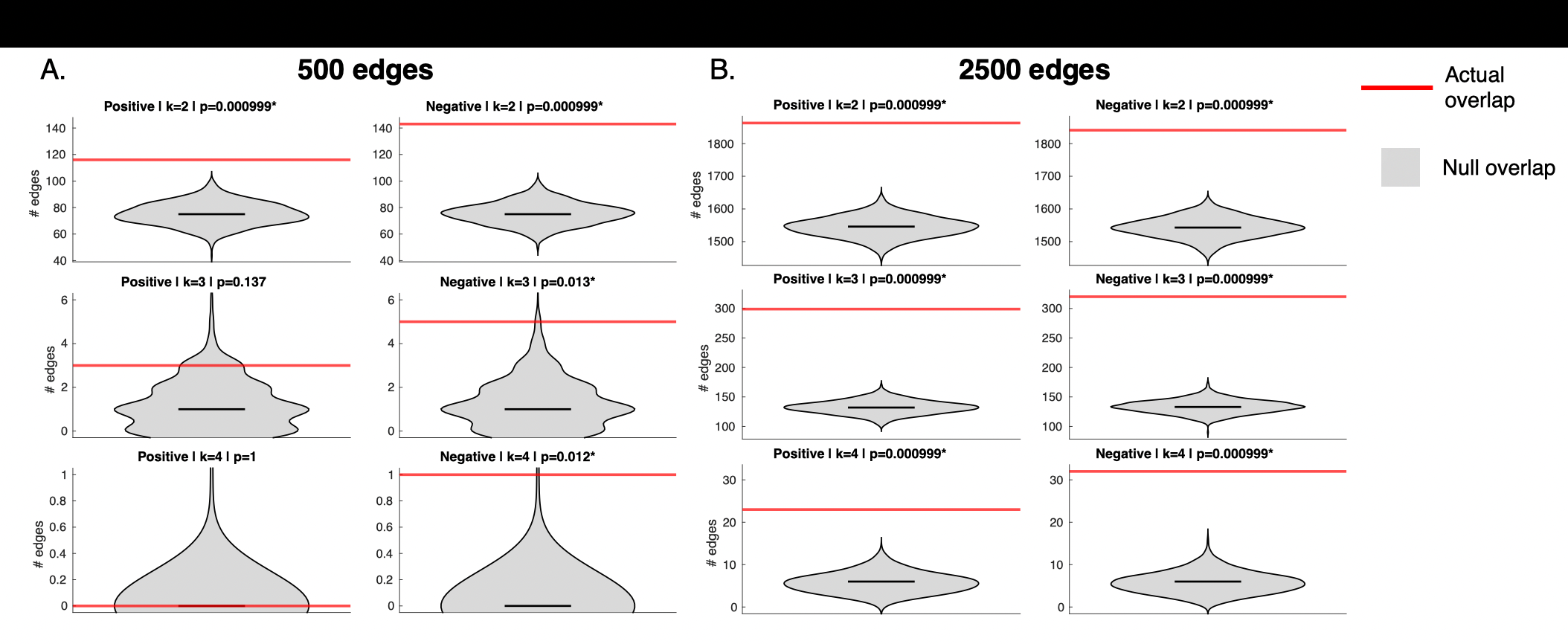

Supplemental Figure 7. Quantifying the occurrence of individual edges across models. Results using A) 500 edges in the positive and negative networks (1,000 total) and B) 2,500 edges in the positive and negative networks (5,000 total). All plots: Violin plots show the number of edges shared across models as a red horizontal line in each plot. The grey violin plot represents the null distribution of the number of shared edges obtained from 1,000 permutations, with the black bar in the violin plot indicating the median of the 1,000 permutations. The positive network is shown in the left column; the negative network, in the right column. Above each plot, ‘k’ corresponds to the edge occurrence across models. *P*-values are shown at the top of each plot; asterisks (*) indicate statistical significance after multiple comparisons correction.

*Edge overlap using different thresholds*

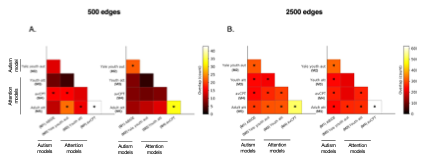

Supplemental Figure 8. Pairwise edge overlap between models. A) Results using 500 edges in each network (1,000 total) and B) 2,500 edges in each network (5,000 total). For both panels, two matrices are shown: overlap between the highest positively ranked edges (‘Pos-Pos’, left); overlap between highest negatively ranked edges (‘Neg-Neg’, right). Each cell corresponds to the number of edges shared between models. Dataset from which the model was obtained is indicated along the rows and columns of each matrix. The colorbar is scaled so that fewer shared edges (in terms of absolute number) are darker colors; more shared edges are lighter colors. An asterisk indicates a statistically significant number of shared edges between the two models after correcting for multiple comparisons. ABIDE, autism brain imaging data exchange. Att, attention; Aut, autism; avCPT, audio-visual continuous performance task; M, model; Neg net, negative network; Pos net, positive network.

| **Model** | **Condition** | **Rank** | **Node** | **Degree** | **MNI (x, y, z)** | **Hemisphere, lobe** |
| --- | --- | --- | --- | --- | --- | --- |
| Positive | Shared by 2 models | 1 | 238 | 18 | -36.95, -52.94, -31.11 | L-Cerebellum |
|  |  | 2 | 78 | 17 | 23.7, -96.01, 6.45 | R-Occipital |
|  |  | 3 | 212 | 12 | -10.83, -98.14, 7.63 | L-Occipital |
|  |  | 4 | 208 | 10 | -16.79, -84.92, 33.04 | L-Occipital |
|  |  | 5 | 171 | 10 | -50.64, -23.79, 41.37 | L-Parietal |
|  |  | 6 | 165 | 10 | -45.8, -0.43, 49.28 | L-Motor strip |
|  |  | 7 | 159 | 10 | -58.09, -5.56, 27.19 | L-Motor strip |
|  |  | 8 | 140 | 10 | -5.96, 48.09, 11.72 | L-Prefrontal |
|  |  | 9 | 76 | 10 | 17.94, -83.43, -11.28 | R-Occipital |
|  |  | 10 | 64 | 10 | 56.49, -8.54, -14.27 | R-Temporal |
|  |  | 11 | 21 | 10 | 55.35, 9.62, 22.22 | R-Prefrontal |
|  |  | 12 | 200 | 9 | -42.57, -52.11, -17.36 | L-Temporal |
|  |  | 13 | 44 | 9 | 7.48, -57.28, 61.79 | R-Parietal |
|  |  | 14 | 23 | 9 | 57.78, -8.34, 27.32 | R-Motor strip |
|  |  | 15 | 216 | 8 | -22.11, -66.7, 7.45 | L-Occipital |
| Positive | Shared by 3 models | 1 | 238 | 3 | -36.95, -52.94, -31.11 | L-Cerebellum |
|  |  | 2 | 141 | 3 | -11.7, 65.09, 4.18 | L-Prefrontal |
|  |  | 3 | 128 | 3 | 5.46, -9.67, 5.24 | R-Subcortical |
|  |  | 4 | 98 | 3 | 14.6, -46.08, 2.84 | R-Limbic |
|  |  | 5 | 212 | 2 | -10.83, -98.14, 7.63 | L-Occipital |
|  |  | 6 | 208 | 2 | -16.79, -84.92, 33.04 | L-Occipital |
|  |  | 7 | 166 | 2 | -27.58, -9.08, 55.86 | L-Motor strip |
|  |  | 8 | 158 | 2 | -41.59, -14.68, 44.79 | L-Motor strip |
|  |  | 9 | 94 | 2 | 35.59, -14.71, -18.45 | R-Limbic |
|  |  | 10 | 78 | 2 | 23.7, -96.01, 6.45 | R-Occipital |
|  |  | 11 | 50 | 2 | 48.95, -58.12, 14.36 | R-Parietal |
|  |  | 12 | 32 | 2 | 32.05, -5.36, 52.05 | R-Motor strip |
|  |  | 13 | 264 | 1 | -11.61, -25.62, 14.8 | L-Subcortical |
|  |  | 14 | 259 | 1 | -10.4, 10.9, -8.13 | L-Subcortical |
|  |  | 15 | 256 | 1 | -24.3, -37.79, -44.28 | L-Cerebellum |
| Positive | Shared by at least 2 models | 1 | 238 | 21 | -36.95, -52.94, -31.11 | L-Cerebellum |
|  |  | 2 | 78 | 19 | 23.7, -96.01, 6.45 | R-Occipital |
|  |  | 3 | 212 | 14 | -10.83, -98.14, 7.63 | L-Occipital |
|  |  | 4 | 208 | 12 | -16.79, -84.92, 33.04 | L-Occipital |
|  |  | 5 | 171 | 11 | -50.64, -23.79, 41.37 | L-Parietal |
|  |  | 6 | 165 | 11 | -45.8, -0.43, 49.28 | L-Motor strip |
|  |  | 7 | 141 | 11 | -11.7, 65.09, 4.18 | L-Prefrontal |
|  |  | 8 | 98 | 11 | 14.6, -46.08, 2.84 | R-Limbic |
|  |  | 9 | 166 | 10 | -27.58, -9.08, 55.86 | L-Motor strip |
|  |  | 10 | 159 | 10 | -58.09, -5.56, 27.19 | L-Motor strip |
|  |  | 11 | 140 | 10 | -5.96, 48.09, 11.72 | L-Prefrontal |
|  |  | 12 | 128 | 10 | 5.46, -9.67, 5.24 | R-Subcortical |
|  |  | 13 | 76 | 10 | 17.94, -83.43, -11.28 | R-Occipital |
|  |  | 14 | 64 | 10 | 56.49, -8.54, -14.27 | R-Temporal |
|  |  | 15 | 23 | 10 | 57.78, -8.34, 27.32 | R-Motor strip |
| Negative | Shared by 2 models | 1 | 238 | 22 | -36.95, -52.94, -31.11 | L-Cerebellum |
|  |  | 2 | 187 | 15 | -49.49, 11.11, -30.56 | L-Temporal |
|  |  | 3 | 21 | 15 | 55.35, 9.62, 22.22 | R-Prefrontal |
|  |  | 4 | 78 | 14 | 23.7, -96.01, 6.45 | R-Occipital |
|  |  | 5 | 64 | 13 | 56.49, -8.54, -14.27 | R-Temporal |
|  |  | 6 | 265 | 11 | -4.97, -21.51, -15.83 | L-Brainstem |
|  |  | 7 | 193 | 11 | -59.85, -27.42, -18.14 | L-Temporal |
|  |  | 8 | 113 | 11 | 36.7, -57.1, -32.77 | R-Cerebellum |
|  |  | 9 | 128 | 10 | 5.46, -9.67, 5.24 | R-Subcortical |
|  |  | 10 | 59 | 10 | 43.36, -26.48, -24.63 | R-Temporal |
|  |  | 11 | 53 | 10 | 52.84, 10.9, -21.83 | R-Temporal |
|  |  | 12 | 50 | 10 | 48.95, -58.12, 14.36 | R-Parietal |
|  |  | 13 | 263 | 9 | -4.87, -10.34, 5.83 | L-Subcortical |
|  |  | 14 | 173 | 9 | -41.23, -15.57, 14.45 | L-Parietal |
|  |  | 15 | 242 | 8 | -30.17, -80.21, -40.35 | L-Cerebellum |
| Negative | Shared by 3 models | 1 | 213 | 4 | -14.72, -83.97, -13.06 | L-Occipital |
|  |  | 2 | 200 | 3 | -42.57, -52.11, -17.36 | L-Temporal |
|  |  | 3 | 21 | 3 | 55.35, 9.62, 22.22 | R-Prefrontal |
|  |  | 4 | 263 | 2 | -4.87, -10.34, 5.83 | L-Subcortical |
|  |  | 5 | 244 | 2 | -6.5, -50.18, -11.37 | L-Cerebellum |
|  |  | 6 | 222 | 2 | -8.6, -58.79, 17.61 | L-Limbic |
|  |  | 7 | 148 | 2 | -11.17, 34.26, 51.48 | L-Prefrontal |
|  |  | 8 | 145 | 2 | -10.15, 55.69, 30.24 | L-Prefrontal |
|  |  | 9 | 106 | 2 | 7.15, -68.97, -20.89 | R-Cerebellum |
|  |  | 10 | 105 | 2 | 6.92, -67.95, -37.77 | R-Cerebellum |
|  |  | 11 | 88 | 2 | 7.01, -18.84, 29.78 | R-Limbic |
|  |  | 12 | 85 | 2 | 5.12, -38.94, 27.02 | R-Limbic |
|  |  | 13 | 76 | 2 | 17.94, -83.43, -11.28 | R-Occipital |
|  |  | 14 | 72 | 2 | 20.97, -63.69, -9 | R-Occipital |
|  |  | 15 | 65 | 2 | 59.25, -43.77, 8.46 | R-Temporal |
| Negative | Shared by 4 models | 1 | 200 | 1 | -42.57, -52.11, -17.36 | L-Temporal |
|  |  | 2 | 145 | 1 | -10.15, 55.69, 30.24 | L-Prefrontal |
| Negative | Shared by at least 2 models | 1 | 238 | 22 | -36.95, -52.94, -31.11 | L-Cerebellum |
|  |  | 2 | 21 | 18 | 55.35, 9.62, 22.22 | R-Prefrontal |
|  |  | 3 | 187 | 15 | -49.49, 11.11, -30.56 | L-Temporal |
|  |  | 4 | 78 | 14 | 23.7, -96.01, 6.45 | R-Occipital |
|  |  | 5 | 64 | 14 | 56.49, -8.54, -14.27 | R-Temporal |
|  |  | 6 | 113 | 12 | 36.7, -57.1, -32.77 | R-Cerebellum |
|  |  | 7 | 265 | 11 | -4.97, -21.51, -15.83 | L-Brainstem |
|  |  | 8 | 263 | 11 | -4.87, -10.34, 5.83 | L-Subcortical |
|  |  | 9 | 200 | 11 | -42.57, -52.11, -17.36 | L-Temporal |
|  |  | 10 | 193 | 11 | -59.85, -27.42, -18.14 | L-Temporal |
|  |  | 11 | 59 | 11 | 43.36, -26.48, -24.63 | R-Temporal |
|  |  | 12 | 53 | 11 | 52.84, 10.9, -21.83 | R-Temporal |
|  |  | 13 | 145 | 10 | -10.15, 55.69, 30.24 | L-Prefrontal |
|  |  | 14 | 128 | 10 | 5.46, -9.67, 5.24 | R-Subcortical |
|  |  | 15 | 50 | 10 | 48.95, -58.12, 14.36 | R-Parietal |
| Both networks | Shared by at least 2 models in either positive or negative | 1 | 238 | 43 | -36.95, -52.94, -31.11 | L-Cerebellum |
|  |  | 2 | 78 | 33 | 23.7, -96.01, 6.45 | R-Occipital |
|  |  | 3 | 21 | 28 | 55.35, 9.62, 22.22 | R-Prefrontal |
|  |  | 4 | 64 | 24 | 56.49, -8.54, -14.27 | R-Temporal |
|  |  | 5 | 200 | 20 | -42.57, -52.11, -17.36 | L-Temporal |
|  |  | 6 | 128 | 20 | 5.46, -9.67, 5.24 | R-Subcortical |
|  |  | 7 | 212 | 19 | -10.83, -98.14, 7.63 | L-Occipital |
|  |  | 8 | 50 | 19 | 48.95, -58.12, 14.36 | R-Parietal |
|  |  | 9 | 263 | 18 | -4.87, -10.34, 5.83 | L-Subcortical |
|  |  | 10 | 171 | 18 | -50.64, -23.79, 41.37 | L-Parietal |
|  |  | 11 | 187 | 17 | -49.49, 11.11, -30.56 | L-Temporal |
|  |  | 12 | 163 | 17 | -56.98, -3.43, 6.82 | L-Motor strip |
|  |  | 13 | 141 | 17 | -11.7, 65.09, 4.18 | L-Prefrontal |
|  |  | 14 | 140 | 17 | -5.96, 48.09, 11.72 | L-Prefrontal |
|  |  | 15 | 53 | 17 | 52.84, 10.9, -21.83 | R-Temporal |

Supplemental Table 4. Highest degree nodes shared across models. Shown are the top 15 nodes in terms of degree (the number of edges connected to that node). Note that the positive network model does not share edges in four models, and the negative network model only shared one edge across four models, so only two nodes are shown. L, left; MNI, Montreal Neurological Institute coordinate space; R, right.

*Visualization of edges when models predicted congruent phenotypes: autism models*

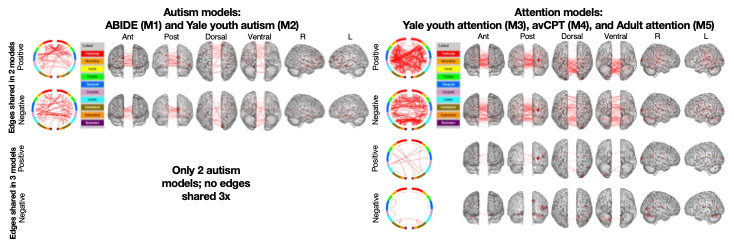

Supplemental Figure 9. Determining the anatomical location of shared edges in the positive and negative network models when autism traits are predicted. Top panel: Positive network masks. Bottom panel: negative network masks. A circle plot is shown immediately to the right of the label indicating the number of times an edge is shared. The topmost aspect of the circle represents anterior; the bottom, posterior. The right half of the circle plot corresponds to the right hemisphere of the brain. A legend indicating the approximate anatomic “lobe” is shown to the top right of the circle plots. The same edges are plotted in the glass brains as lines connecting different nodes; in these visualizations, nodes are sized according to degree, the number of edges connected to that node. The view of the brain is indicated at the top of the panel. ABIDE, autism brain imaging data exchange; Ant, anterior; L, left; M, model; Post, posterior; R, right.

*Visualization of edges when models predicted congruent phenotypes: attention models*

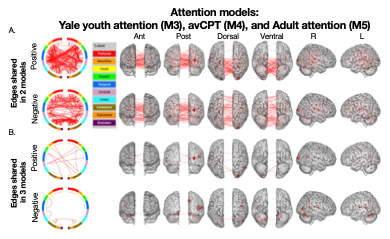

Supplemental Figure 10. Determining the anatomical location of shared edges in the positive and negative network models when sustained attention performance is predicted. A) Results when edges are shared in two models. B) Results when edges are shared in three models. For both A) and B), the top panel shows positive network masks; in the bottom panel, negative network masks. A circle plot is shown immediately to the right of the label indicating the number of times an edge is shared. The topmost aspect of the circle represents anterior; the bottom, posterior. The right half of the circle plot corresponds to the right hemisphere of the brain. A legend indicating the approximate anatomic “lobe” is shown to the top right of the circle plots. The same edges are plotted in the glass brains as lines connecting different nodes; in these visualizations, nodes are sized according to degree, the number of edges connected to that node. The view of the brain is indicated at the top of the panel. Ant, anterior; avCPT, audio-visual continuous performance task; L, left; M, model; Post, posterior; R, right.

*Network matrices using different thresholds*

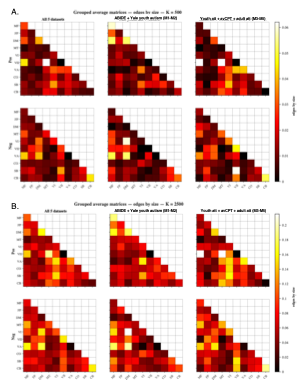

Supplemental Figure 11. Neuroanatomy of the network models using different thresholds. A) Results using 500 edges. B) Results using 2,500 edges. For all plots, the leftmost column corresponds to a summary matrix for all five datasets. Middle column corresponds to a summary matrix for the datasets predicting autistic phenotypes. Rightmost column corresponds to a summary matrix for the datasets predicting continuous attention task performance. For each edge threshold, the positive network masks are shown in the top row; negative network masks, in the bottom row. Each cell in a matrix corresponds to the fraction of edges in a given network pair that were included in the given model, after correcting for network size. The colorbars for each matrix are scaled so that a lower fraction of edges corresponds to darker colors; a higher fraction of edges corresponds to lighter colors. The title of the colorbar ‘edges by size’ corresponds to the fraction of edges in a cell. Statistically significant network pairs after multiple comparisons correction are denoted with an asterisk (*). ABIDE, autism brain imaging data exchange. Att, attention; Aut, autism; avCPT, audio-visual continuous performance task; Neg, negative network; Pos, positive network. Network labels: MF, medial frontal; FP, frontoparietal; DM, default mode; MT, motor; VI, visual I; VII, visual II; VA, visual association; CO, cingulo-opercular; SB, subcortical; CB, cerebellum.

*Neuroanatomy of the network models: individual datasets and pairwise relationships*

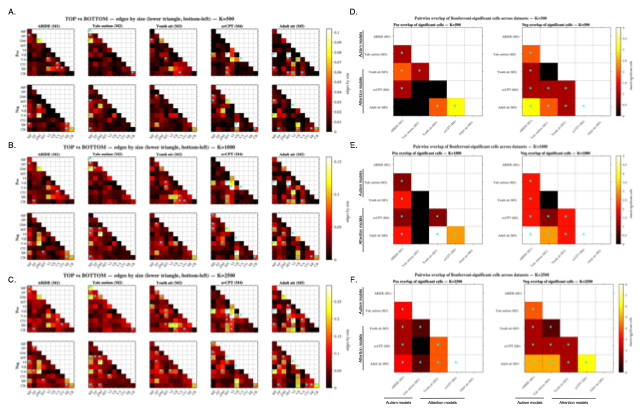

Supplemental Figure 12. Neuroanatomy of the network models: individual datasets and pairwise relationships across different edge selection thresholds. A-C) Results of individual datasets at different thresholds; the number of edges used in each threshold (A = 500; B = 1,000; C = 2,500) is indicated above each set of matrices. For each edge threshold, the positive network masks are shown in the top row; negative network masks, in the bottom row. Each cell in a matrix corresponds to the fraction of edges in a given network pair that were included in the given model, after correcting for network size. The colorbars for each matrix are scaled so that a lower fraction of edges corresponds to darker colors; a higher fraction of edges corresponds to lighter colors. The title of the colorbar ‘edges by size’ corresponds to the fraction of edges in a cell. In all rows, the first two columns correspond to models predictive of autistic phenotypes; columns 3-5 correspond to models predictive of continuous attention phenotypes. D-F) Assessing pairwise relationships of significant network pairs. The edge threshold is indicated above each set of matrices. The left column corresponds to the positive network; the right column, the negative network. The colorbars for each matrix are scaled so that a lower number of shared network pairs corresponds to darker colors; a higher number of shared network pairs corresponds to lighter colors. For all plots, statistically significant network pairs after multiple comparisons correction are denoted with an asterisk (*). Bonferroni correction was applied to the data in A-C (correcting for 55 tests per matrix) and to the data in D-F (correcting for 10 tests per matrix). ABIDE, autism brain imaging data exchange. Att, attention; Aut, autism; avCPT, audio-visual continuous performance task; M, model; Neg, negative network; Pos, positive network. Network labels: MF, medial frontal; FP, frontoparietal; DM, default mode; MT, motor; VI, visual I; VII, visual II; VA, visual association; CO, cingulo-opercular; SB, subcortical; CB, cerebellum.

|  | Auditory avCPT condition | |  | | | |
| --- | --- | --- | --- | --- | --- | --- |
|  |  | Autism models | | Attention models | | |
|  |  | ABIDE (M1) | Yale youth autism (M2) | Youth attention (M3) | avCPT (M4) | Adult attention (M5) |
| Autism models | ABIDE (M1) | — | 0.132* | -0.048 | -0.018 | -0.072 |
|  | Yale youth autism (M2) | 0.132* | — | -0.095 | -0.082* | -0.148* |
| Attention models | Youth attention (M3) | -0.048 | -0.095 | — | 0.057 | 0.149* |
|  | avCPT (M4) | -0.018 | -0.082* | 0.057 | — | 0.297* |
|  | Adult attention (M5) | -0.072 | -0.148* | 0.149* | 0.297* | — |
|  | Visual avCPT condition | |  | | | |
|  |  | Autism models | | Attention models | | |
|  |  | ABIDE (M1) | Yale youth autism (M2) | Youth attention (M3) | avCPT (M4) | Adult attention (M5) |
| Autism models | ABIDE (M1) | — | 0.141* | -0.056 | -0.063 | -0.077 |
|  | Yale youth autism (M2) | 0.141* | — | -0.09 | -0.083 | -0.144* |
| Attention models | Youth attention (M3) | -0.056 | -0.09 | — | 0.140* | 0.146* |
|  | avCPT (M4) | -0.063 | -0.083 | 0.140* | — | 0.376* |
|  | Adult attention (M5) | -0.077 | -0.144* | 0.146* | 0.376* | — |

Supplemental Table 5. A similarity matrix (in table format) showing similarity of edge-behavior vectors across datasets. Top) Results using the avCPT auditory condition. Bottom) Results using the avCPT visual condition. Shown in each cell is the Pearson correlation coefficient. Asterisks (*) indicate statistical significance after correction for multiple comparisons. ABIDE, autism brain imaging data exchange dataset; avCPT, audio-visual continuous performance task dataset, M, model

Horien, C., Mandino, F., Greene, A.S. *et al.* Optimizing functional connectivity scanning conditions for predicting autistic traits. *Nat. Mental Health* (2026).

Horien C, Noble S, Finn ES, Shen X, Scheinost D, Constable RT. 2018. Considering factors affecting the connectome-based identification process: Comment on Waller et al. Neuroimage. 169:172-175.

Horien C, Shen XL, Scheinost D, Constable RT. 2019. The individual functional connectome is unique and stable over months to years. Neuroimage. 189:676-687.

Ingersoll B. 2010. Broader autism phenotype and nonverbal sensitivity: evidence for an association in the general population. J Autism Dev Disord. 40:590-598.

Joshi A, Scheinost D, Okuda H, Belhachemi D, Murphy I, Staib LH, Papademetris X. 2011. Unified framework for development, deployment and robust testing of neuroimaging algorithms. Neuroinformatics. 9:69-84.

Ju Y, Horien C, Chen W, Guo W, Lu X, Sun J, Dong Q, Liu B, Liu J, Yan D, Wang M, Zhang L, Guo H, Zhao F, Zhang Y, Shen X, Constable RT, Li L. 2020. Connectome-based models can predict early symptom improvement in major depressive disorder. J Affect Disord. 273:442-452.

Lake EMR, Finn ES, Noble SM, Vanderwal T, Shen X, Rosenberg MD, Spann MN, Chun MM, Scheinost D, Constable RT. 2019. The Functional Brain Organization of an Individual Allows Prediction of Measures of Social Abilities Transdiagnostically in Autism and Attention-Deficit/Hyperactivity Disorder. Biol Psychiatry. 86:315-326.

Lichenstein SD, Scheinost D, Potenza MN, Carroll KM, Yip SW. 2021. Dissociable neural substrates of opioid and cocaine use identified via connectome-based modelling. Mol Psychiatry. 26:4383-4393.

Lord C RM, DiLavore PC, Risi S, Gotham K, Bishop S. 2012. Autism Diagnostic Observation Schedule, Second Edition. Torrance, CA: Western Psychological Services.

Rapuano KM, Rosenberg MD, Maza MT, Dennis NJ, Dorji M, Greene AS, Horien C, Scheinost D, Todd Constable R, Casey BJ. 2020. Behavioral and brain signatures of substance use vulnerability in childhood. Dev Cogn Neurosci. 46:100878.

Rosenberg M, Noonan S, DeGutis J, Esterman M. 2013. Sustaining visual attention in the face of distraction: a novel gradual-onset continuous performance task. Atten Percept Psycho. 75:426-439.

Rosenberg MD, Finn ES, Scheinost D, Papademetris X, Shen X, Constable RT, Chun MM. 2016. A neuromarker of sustained attention from whole-brain functional connectivity. Nat Neurosci. 19:165-171.

Rosenberg MD, Scheinost D, Greene AS, Avery EW, Kwon YH, Finn ES, Ramani R, Qiu M, Constable RT, Chun MM. 2020. Functional connectivity predicts changes in attention observed across minutes, days, and months. Proc Natl Acad Sci U S A. 117:3797-3807.

Satterthwaite TD, Elliott MA, Gerraty RT, Ruparel K, Loughead J, Calkins ME, Eickhoff SB, Hakonarson H, Gur RC, Gur RE, Wolf DH. 2013. An improved framework for confound regression and filtering for control of motion artifact in the preprocessing of resting-state functional connectivity data. Neuroimage. 64:240-256.

Yip SW, Scheinost D, Potenza MN, Carroll KM. 2019. Connectome-Based Prediction of Cocaine Abstinence. Am J Psychiatry. 176:156-164.
